# Modeling the Anticipated Public Health Benefits of the Next-Generation COVID-19 mRNA-1283 vaccine: An Interim U.S. Population-Level Impact Assessment

**DOI:** 10.1101/2025.09.26.25336704

**Authors:** Kelly Fust, Michele Kohli, Keya Joshi, Shannon Cartier, Amy Lee, Nicolas Van de Velde, Milton Weinstein, Ekkehard Beck

**Author notes:** Address for Correspondence: Ekkehard Beck.

## Abstract

**Aims:** COVID-19 disease burden in United States (US) older adults ≥65 years and persons with underlying medical conditions remains high. This modeling study provides an interim estimate of the anticipated public health impact of the next-generation COVID-19 mRNA-1283 vaccine in these populations at high-risk of severe COVID-19 outcomes.

**Methods:** mRNA-1283 was compared to no vaccination and originally licensed mRNA COVID-19 vaccines mRNA-1273 and BNT162b2. Analyses were conducted using a static decision-analytic model (1-year horizon). Vaccine effectiveness (VE) against infection and hospitalization for mRNA-1283 versus no vaccination was based on the relative VE (rVE) from the Phase 3 pivotal randomized controlled trial comparing mRNA-1283 against mRNA-1273 and mRNA-1273 real-world data. rVE estimates for mRNA-1283 versus BNT162b2 were based on an indirect treatment comparison. Clinical outcomes calculated included total numbers of symptomatic infections, outpatient and long COVID cases, hospitalizations, and deaths. Sensitivity and scenario analyses were performed.

**Results:** During the 2024/2025 season in the US, a single dose of the mRNA-1283 vaccine was estimated to prevent approximately 2.9 (1.3-4.3) million symptomatic infections, 171,000 (77,000-260,000) hospitalizations, and 22,350 (10,050-33,480) deaths compared to no vaccination. Compared to BNT162b2, mRNA-1283 was estimated to avert an additional 0.79 million symptomatic infections, 58,000 hospitalizations, and 7,565 deaths. Compared to mRNA-1273, mRNA-1283 was estimated to avert an additional 0.56 million symptomatic infections, 46,000 hospitalizations, and 5,920 deaths. Across all scenarios the majority of severe COVID-19 cases (i.e., hospitalizations and deaths) were prevented among older adults ≥65 years.

**Limitations:** The real-world effectiveness and safety of mRNA-1283 have not yet been established and the relative VE estimates should be validated with real-world data. Future COVID-19 incidence and incidence pattern throughout the season is uncertain.

**Conclusions:** Interim results suggest that the next-generation COVID-19 mRNA-1283 vaccine could substantially reduce the clinical burden of COVID-19 among those at high risk of severe disease. Compared to no vaccination and originally approved mRNA vaccines, mRNA-1283 provides a valuable option to potentially enhance COVID-19 immunization programmes and protection of those most vulnerable.

## Introduction

Although the pandemic has ended, COVID-19 continues to place a significant burden on the United States (US) healthcare system. The Centers for Disease Control and Prevention (CDC) estimates that there were between 10.1 to 16.4 million COVID-19 illnesses, 280,000–450,000 COVID-19 hospitalizations and 23,000–52,000 COVID-19 deaths, between October 2024 and June 2025^1^. During this timeframe, COVID-19 hospitalizations were primarily in older adults, with cumulative hospitalization rates (per 100,000) of 317.4 in those aged 65 years and over, compared with 61.2 in those aged 50–64, and 20.5 in those aged 18–49 years. In addition to older adults, patients with underlying medical conditions are at increased risk of severe COVID-19 disease. Based on US CDC data presented at the Advisory Committee on Immunization Practices (ACIP) meeting in April 2025, 18 to 49 year old patients with diabetes or coronary artery disease were observed to have a 9 and 8 times higher hospitalization rate, respectively, than the overall population of community dwelling adults in the US^2^. Despite the well-documented increased risks in adults over 65 years of age, uptake of the updated 2024/25 COVID-19 vaccine only reached 44.4% by April 5, 2025^3^, which was lower than the 2024/25 influenza vaccine uptake in the same age group.^4^

mRNA-1283 (mNEXSPIKE, Moderna) is a FDA licensed^5^, next generation COVID-19 vaccine with an innovative design, developed to focus the immune responses to potentially increase protection against COVID-19 and increased product stability compared with the originally licensed mRNA COVID-19 vaccines such as mRNA-1273^6^ (Spikevax, Moderna) or BNT162b2 (Comirnaty, Pfizer-BioNTech). In comparison to mRNA-1273 and BNT162b2, which encode the full-length SARS-CoV-2 spike protein, the next-generation COVID-19 vaccine mRNA-1283 uses an innovative approach to focus only on the receptor binding domain (RBD) and N-terminal domain (NTD) of the SARS-CoV-2 spike protein. Both of these domains contain immunodominant epitopes for neutralizing antibodies against SARS-CoV-2 and allowing enhanced immune response at a lower mRNA dose of 10 µg (1/5th of dose of mRNA-1273)^6^. Additionally, shorter mRNA sequences are more stable ^7^ and may therefore improve refrigeration stability^8^.

The pivotal NextCOVE randomized observer-blind active controlled phase 3 trial demonstrated the efficacy of mRNA-1283 compared to mRNA-1273^6^. Individuals aged 12 years and over were randomized to receive a bivalent original and Omicron-containing BA.4/5 formulation of mRNA-1283 or mRNA-1273 between March 23 and August 23, 2023. Higher immunogenicity of mRNA-1283 over mRNA-1273 was observed with a geometric mean titer ratio (GMR) against BA.4/5 of 1.3 (95% CI 1.2-1.5) among the overall population and an increased GMR of 1.8 (95% CI 1.4-2.2) among the vulnerable group of 65 years and older. The relative vaccine efficacy (rVE) of mRNA-1283 versus mRNA-1273 was 9.3% (99.4 % CI --6.6–22.8%) for COVID-19 symptomatic infection in the overall population, with greater protective benefit against infection in those aged 65 years and over (13.5%, 95% CI -7.7–30.6%)^6^ and in those with underlying medical conditions putting them at higher risk of severe COVID-19 disease (≥12 years 16.3%, 95% CI 1.8-28.7%; ≥65 years 28.1%, 95% CI 4.4-45.9%)^6^. In a post-hoc analysis, the rVE of mRNA-1283 vs mRNA-1273 against FDA defined severe COVID-19 was estimated at 38.1% (95% CI -6.7–64.1%)^6^.

A substantial body of real-world evidence suggests that mRNA-1273 is more protective than BNT162b2. This evidence includes head-to-head real-world effectiveness studies and GRADE meta-analyses of pairwise studies which capture different vaccine versions up until bivalent vaccines containing the original strain and Omicron BA.4/5. For example, in a large real-world study comparing the effectiveness of mRNA-1273 and BNT162b2 bivalent original/BA.4/ 5 COVID-19 vaccines in adults during the 2022/23 season, the relative VE (rVE) against outpatient visits and hospitalization of mRNA-1273 was higher by 5.1% (95% CI 3.2–6.9%) and 9.8% (95% CI 2.6–16.4%), respectively, compared with BNT162b2^9^. This enhanced protective effect was also seen in older adults and adults with underlying medical conditions. A systematic literature review (SLR) and meta-analysis (MA) reported a lower risk of symptomatic infections (risk ratio [RR] 0.74, 95% CI 0.56–0.97) and hospitalizations (RR 0.69, 95% CI 0.53–0.89) with mRNA-1273 compared with BNT162b2 in those 65 years and over^10^. Another SLR and MA focused on adults with medical conditions also found mRNA-1273 was more protective against infections (RR 0.85, 95% CI 0.79–0.92) and hospitalizations (RR 0.88, 95% CI 0.82–0.94) compared with BNT162b2, in adults with at least one underlying medical condition^11^. Based on these real-world effectiveness (RWE) data, mRNA-1273 may likely deliver greater protection against infection and hospitalizations than BNT162b2. Phase 3 RCT data suggest that mRNA-1283 provides greater protection compared with mRNA-1273 and therefore it is expected that mRNA-1283 can provide greater protection than BNT162b2. An indirect treatment comparison (ITC) established mRNA-1283 to be more effective than BNT162b2^12^, using data from the NextCOVE trial^6^ and a large real-world study comparing mRNA-1273 vs BNT162b2 in adults aged 18 and over^9^. The rVE against symptomatic COVID-19 infection of mRNA-1283 compared with BNT162b2 was 15.3% (95% CI 4.7-24.8%) among all adults and even more pronounced in those at highest unmet need such as adults 65 years and older with an rVE of 22.8% (95% CI 4.7–24.8%)^9^.

In May 2025, the Food and Drug Administration (FDA) authorized the use of mRNA-1283 for those 65 years of age and older, and 12 years through 64 years of age with at least one underlying condition that puts them at high risk for severe outcomes from COVID-19^11^. As the COVID-19 disease burden in the US remains high, especially for those at high risk for severe COVID-19 disease, decision makers are faced with the challenge to optimize the COVID-19 vaccination policy. Although the 2025/2026 U.S. CDC recommendation for COVID-19 vaccination is still pending, this modeling study was conducted to provide an interim estimate of the anticipated public health impact of the next-generation mRNA-1283 vaccine. The analysis compares mRNA-1283 to no vaccination and to originally licensed mRNA vaccines (mRNA-1273 and BNT162b2) within the licensed target population for mRNA-1283 in the US.

## Methods

### Overview

A Markov model with monthly cycles was used to project the potential number of symptomatic COVID-19 infections and associated COVID-19 outcomes (hospitalizations, deaths and cases of long COVID) expected in the target population in the US over a one-year time horizon with a single dose of mRNA-1283 (annual vaccination) compared to no vaccination. A scenario analysis was conducted where those 65 years and older were eligible to receive a second dose a minimum of two months after the first dose (semi-annual vaccination). In addition, vaccination (annual or semi-annual) with mRNA-1283 was compared to vaccination (annual or semi-annual) with mRNA-1273 and BNT162b2^13^.

### Model Structure

The cohort begins the model in the Well health state (Figure 1) in which each person is susceptible to symptomatic COVID-19 infection irrespective of their previous history of COVID-19 infections and vaccinations. In each month moving forward, a portion of those in the Well state may receive a vaccine and are then subject to vaccine-related adverse events (AEs) (AE rates are described in the Technical Appendix; however, these are not reported in this public health impact analysis). Each month, individuals in the Well health state are at risk of symptomatic COVID-19 infection. If they develop an infection, they move through the COVID-19 consequences decision tree (Figure 2) which estimates the subsequent outcomes of infection. Those who die following their COVID-19 infection move into the Dead health state while all others return to the Well health state. Those who did not develop an infection remain in the Well health state.

**Figure 1.**
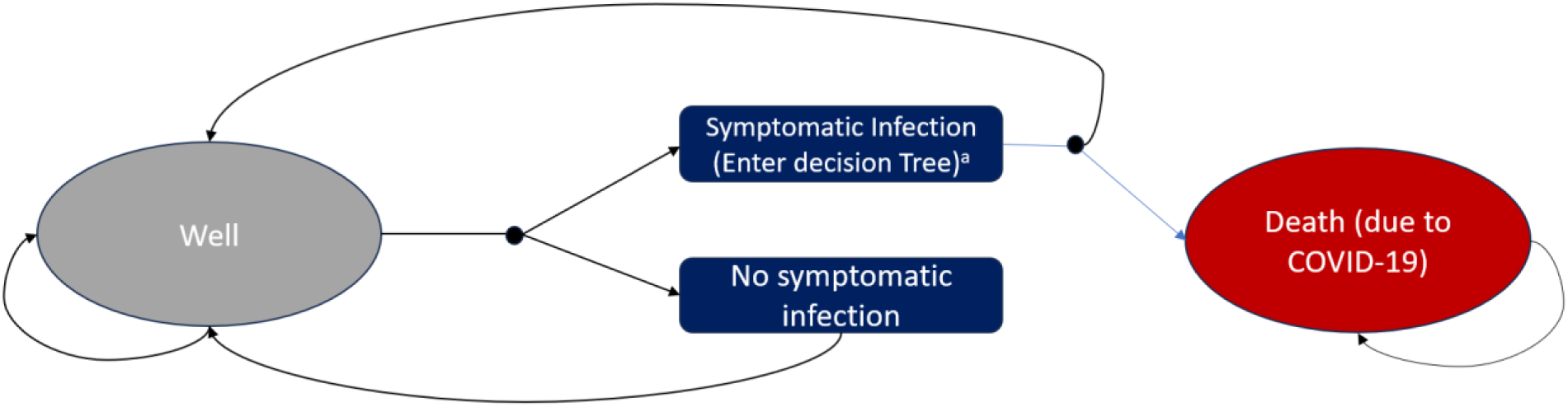
Illustration of the Markov model. ^a^see Figure 2

**Figure 2:**
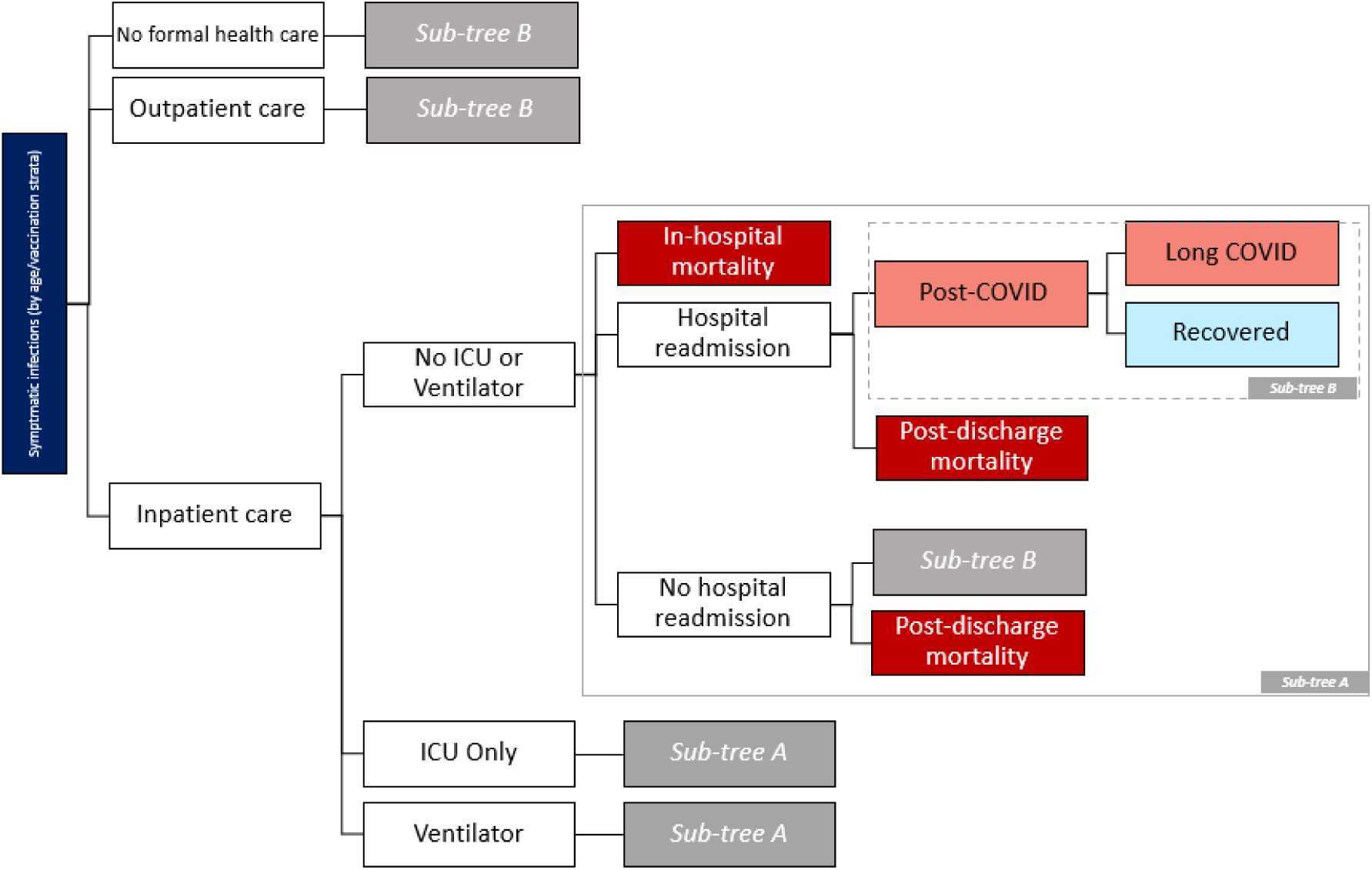
Illustration of the consequences of COVID-19 decision tree structure. ICU: Intensive Care Unit - Not shown in figure: all symptomatic infections are at risk of infection-related myocarditis, and all hospitalizations are followed by post-hospital recovery. An average cost and QALY loss may be assigned for each of these events. - Ventilator refers to mechanical ventilation in an intensive care setting.

The risk of infection and the risk of hospitalization once an infection has developed is reduced in those who are vaccinated. VE declines linearly on a monthly basis within the model. As portion of the cohort can receive a new vaccine each month, the effectiveness calculation is a function of the fraction of the age group vaccinated each month and VE. The hospitalization VE values are adjusted so that the incremental protection against hospitalization above the protection against infection is applied to the probability of hospitalization. (See Technical Appendix).

### Target Population

The size of the cohort, shown in Table 1, was based on the US population^14^. For ages 12-64, the cohort size eligible for vaccination is limited to high-risk individuals, estimated based on data from Kompanieyts et al. (2021)^15^ and Panagiotakopoulos (2025)^16^.

**Table 1:** Population Size.

| Age Group (Years) | Population Size | Percent Eligible* | Eligible Population Size | Source |
| --- | --- | --- | --- | --- |
| 12-17 | 26,766,952 | 25% | 6,691,738 | United Nations (2024)<sup>14</sup>; Kompanieyts et al. (2021)<sup>15</sup> |
| 18-49 | 145,627,454 | 65.7% | 95,738,794 | United Nations (2024)<sup>14</sup>; Panagiotakopoulos (2025)<sup>16</sup> |
| 50-64 | 63,863,350 | 81.0% | 51,729,314 |  |
| ≥65 | 59,874,349 | 100% | 59,874,349 | United Nations (2024)<sup>14</sup> |

\*Reflects the proportion considered high-risk for ages 12-64 years; for ages 65+, all adults are eligible for vaccination.
\*High-risk conditions for severe COVID-19 among those 12-17 years studied by Kompanieyts et al. (2021) include type 1 diabetes, cardiac and circulatory congenital anomalies, obesity, essential hypertension, epilepsy, neuropsychiatric disorders, and asthma as well as children with chronic disease. As Kompanieyts et al. (2021) studied only the prevalence of risk factors among children with a COVID-19 infection but not the prevalence among all children, we assumed a proxy estimate of 25.0% for the prevalence of these risk factors in the current analysis, based on an estimate provided in the introduction section of their study.
<sup>†</sup>High-risk conditions for those 18+ years reflect those included in Panagiotakopoulos et al., and include asthma, cancer (hematologic malignancies), cerebrovascular disease, chronic kidney disease (dialysis), chronic lung diseases (bronchiectasis, COPD, interstitial lung disease, pulmonary embolism, pulmonary hypertension), chronic liver diseases (cirrhosis, non-alcoholic fatty liver disease, alcoholic liver disease, autoimmune hepatitis), cystic fibrosis, diabetes mellitus (types 1 and 2), disabilities, heart conditions, HIV, mental health conditions (depression, schizophrenia spectrum disorders), neurologic conditions (dementia; Parkinson's disease), obesity, physical inactivity, pregnancy and recent pregnancy, primary immunodeficiencies, smoking (current and former), solid organ or blood stem cell transplantation, tuberculosis, use of corticosteroids or other immunosuppressive medications

### Infection Incidence

The model required the incidence of symptomatic COVID-19 infection in the target population when no one receives the vaccine. As these data are not collected by the CDC, this input was approximated based on estimates of observed hospitalization rates per 100,000 by age group from COVID NET^17^, using data on vaccine coverage and effectiveness and the conditional probability of hospitalization given infection in a modified version of the model, as described in the Technical Appendix. Unlike seasonal influenza, COVID-19 hospitalization rates may not only peak during the winter respiratory season but also in the summer, with elevated hospitalization rates observed between these seasonal peaks as well. To estimate the incidence of symptomatic infection, hospitalization rates for one full year, i.e. 12 months, were needed. Thus, the hospitalization rates for September 2023 to August 2024 were used for the base case in this interim analysis of the impact during 2024/2025 season as 12 month 2024/2025 season incidence data at the time of the conduct of the analysis were not available. For high-risk patients, the incidence of symptomatic COVID-19 infection was assumed to be equivalent to the incidence of COVID-19 infection for the general population, as it is assumed that high-risk patients are more likely to experience more severe COVID-19 outcomes but are not at increased risk of developing COVID-19 infection.

### Vaccine Coverage

The vaccine coverage rate (VCR) of the first seasonal dose of the vaccines (annual vaccination) was based on adults aged 18 years and over from COVIDVaxView.^18^ Data from September 2024 to February 2025 were available, with no further uptake assumed after this time (See figure in the Technical Appendix). For the scenario where those aged 65 years and over received a second seasonal dose (semi-annual vaccination), the same coverage as observed during 2023/2024 season was considered (See figure in the Appendix). In absence of CDC data, the VCR for the high-risk population was conservatively assumed to be equivalent to the general population.

### Vaccine Effectiveness

The VE values used for the model analytic time horizon were derived based on the age-specific rVEs of mRNA-1283 compared to the mRNA-1273 vaccine from the pivotal phase 3 randomized clinical trial (RCT) NextCOVE^6,19^ and the 2024-2025 US real-world VEs of mRNA-1273.712 targeting KP.2.^20^ Estimates for high-risk populations in RCT and real-world effectiveness (RWE) studies were used for those aged 18-64 years with high-risk in the model. mRNA-1273 VEs were based on a large, US nationwide RWE study providing interim VE estimates of mRNA-1273. VE against infection and hospitalization were measured at 64 days post administration and were scaled back to 14 days post administration to estimate the initial VE in the model as described in the Technical Appendix. For those aged 12 to 17 years, the 2024-2025 VEs for mRNA-1273 against infection and hospitalization were assumed to be the same and were based on MacNeil’s estimate of the real-world VE of 2024-2025 COVID-19 vaccines against emergency department and urgent care visits in children aged 5-17 years, as these were the only available VE estimates.^21^ These were measured at 81 days in the RWE study. To derive the corresponding initial VE in the model, the observed VE estimate was scaled back 67 days to 14 days post-administration conservatively assuming the monthly VE waning rate against hospitalization^22^. rVE values for mRNA-1283 compared to mRNA-1273 were applied to the mRNA-1273 VEs to calculate the initial VEs for mRNA-1283 as displayed in Table 2. The rVE against COVID-19 CDC defined symptomatic infections^19^ (primary endpoint of Phase 3 RCT NextCOVE) was used as proxy for the rVE against COVID-19 symptomatic infection. The rVE against FDA defined severe COVID-19 (retrospective post-hoc analysis of NextCOVE) was applied as proxy for the rVE against COVID-19 hospitalization. The 95% confidence intervals (CIs) for the VE of mRNA-1283 against infection and hospitalization were derived based on Monte-Carlo simulations considering the 95% CIs of the VE of mRNA-1273 and of the rVE estimates of mRNA-1283 vs mRNA-1273.

**Table 2.** mRNA-1283 COVID-19 Initial Vaccine Effectiveness.

| **Age Group (Years)** | **Symptomatic Infection** |  |  |
| --- | --- | --- | --- |
|  | **2024-2025 mRNA-1273 VE (95% CI)** | **rVE of mRNA-1283 compared to mRNA-1273 (95% CI)** | **mRNA-1283 VE (95% CI)** |
| 12 -17<br>High Risk | 62.4%<br>(38.4%; 77.4%) | 16.3%<br>(1.8%; 28.7%) | 68.5%<br>(50.8%; 85.2%) |
| 18 – 64<br>High Risk | 48.2%<br>(43.3%; 52.7%) | 16.3%<br>(1.8%; 28.7%) | 56.6%<br>(48.5%; 64.4%) |
| >65 | 54.5%<br>(49.5%; 59.0%) | 13.5%<br>(-7.7%; 30.6%) | 60.7%<br>(50.8%; 70.0%) |
|  | **Hospitalization** |  |  |
| 12 -17<br>High Risk | 62.4%<br>(38.4%; 77.4%) | 38.1%<br>(-6.7%; 64.1%) | 76.7%<br>(56.2%; 92.4%) |
| 18 – 64<br>High Risk | 50.5%<br>(29.3%; 65.7%) | 38.1%<br>(-6.7%; 64.1%) | 69.4%<br>(46.3%; 88.2%) |
| >65 | 57.2%<br>(38.3%; 70.8%) | 38.1%<br>(-6.7%; 64.1%) | 73.5%<br>(53.3%; 89.9%) |

CI: Confidence interval; rVE: relative vaccine effectiveness; VE: Vaccine effectiveness

For the scenarios, where mRNA-1283 was compared to mRNA-1273 or BNT-162b2, the initial VE estimates shown in Table 3 were used. To estimate the VE of BNT162b2, the rVE estimates derived in an indirect treatment comparison (ITC)^12,23^ were applied to the VE of mRNA-1283. For the 65 plus and the high-risk 18 to 64 years of age populations the rVE estimate of mRNA-1283 vs BNT162b2 against symptomatic COVID-19 infection of 22.8% (95% CI: 3.7%; 38.1%) and 19.0% (95% CI: 4.9; 31.0%) were assumed. For the rVE against hospitalization, the overall population estimate of 44.1% (95% CI: 3.2; 67.7%) was assumed; this number was derived applying the same methods as outlined in Beck et al. but using the NextCOVE estimate of the rVE of mRNA-1283 vs mRNA-1273 (38.1%, 95% CI: -6.7; 64.1%) against FDA defined severe COVID and the Kopel et al.^24^ estimate of the rVE of mRNA-1273 vs BNT162b2 (9.8%, 95% CI: 2.6; 16.4%) against COVID-19 hospitalization. BNT162b2 VEs against infection and hospitalization were assumed to be the same as mRNA-1273 as the values were based on a mixed population that received either of the two vaccines.

**Table 3.**
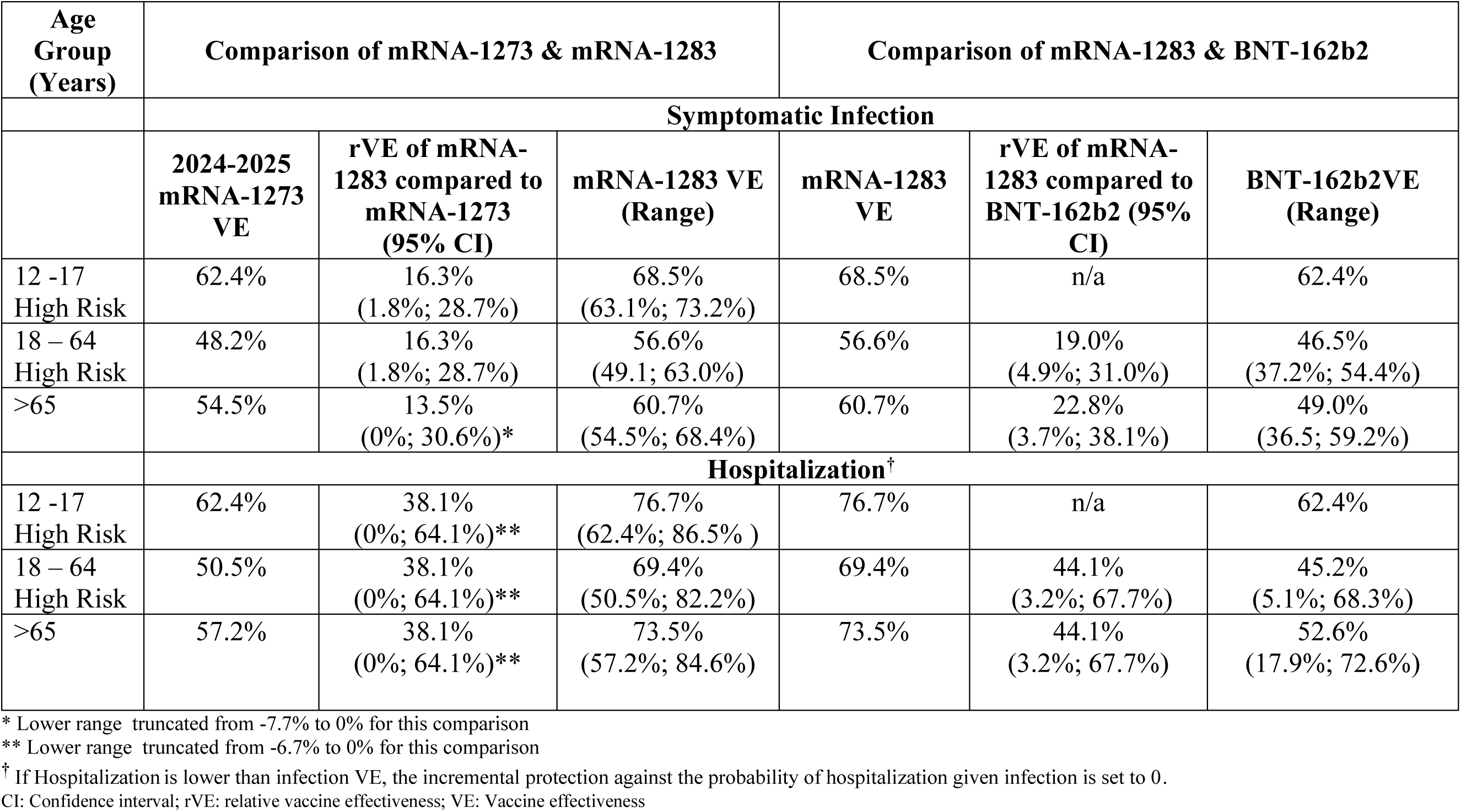
Initial Vaccine Effectiveness for the Comparison of mRNA-1283 to mRNA-1273 and BNT-162b2.

| Age Group (Years) | Comparison of mRNA-1273 & mRNA-1283 |  |  | Comparison of mRNA-1283 & BNT-162b2 |  |  |
| --- | --- | --- | --- | --- | --- | --- |
|  | Symptomatic Infection |  |  |  |  |  |
|  | 2024-2025 mRNA-1273 VE | rVE of mRNA-1283 compared to mRNA-1273 (95% CI) | mRNA-1283 VE (Range) | mRNA-1283 VE | rVE of mRNA-1283 compared to BNT-162b2 (95% CI) | BNT-162b2VE (Range) |
| 12 -17 High Risk | 62.4% | 16.3% (1.8%; 28.7%) | 68.5% (63.1%; 73.2%) | 68.5% | n/a | 62.4% |
| 18 – 64 High Risk | 48.2% | 16.3% (1.8%; 28.7%) | 56.6% (49.1; 63.0%) | 56.6% | 19.0% (4.9%; 31.0%) | 46.5% (37.2%; 54.4%) |
| >65 | 54.5% | 13.5% (0%; 30.6%)* | 60.7% (54.5%; 68.4%) | 60.7% | 22.8% (3.7%; 38.1%) | 49.0% (36.5; 59.2%) |
|  | Hospitalization<sup>†</sup> |  |  |  |  |  |
| 12 -17 High Risk | 62.4% | 38.1% (0%; 64.1%)** | 76.7% (62.4%; 86.5% ) | 76.7% | n/a | 62.4% |
| 18 – 64 High Risk | 50.5% | 38.1% (0%; 64.1%)** | 69.4% (50.5%; 82.2%) | 69.4% | 44.1% (3.2%; 67.7%) | 45.2% (5.1%; 68.3%) |
| >65 | 57.2% | 38.1% (0%; 64.1%)** | 73.5% (57.2%; 84.6%) | 73.5% | 44.1% (3.2%; 67.7%) | 52.6% (17.9%; 72.6%) |

\* Lower range truncated from -7.7% to 0% for this comparison
\*\* Lower range truncated from -6.7% to 0% for this comparison
<sup>†</sup> If Hospitalization is lower than infection VE, the incremental protection against the probability of hospitalization given infection is set to 0.
CI: Confidence interval; rVE: relative vaccine effectiveness; VE: Vaccine effectiveness

The monthly VE waning of mRNA vaccines against infection for Omicron variants relative to those who had never been vaccinated was estimated in a meta-analysis by Higdon et al., (2022)^25^ to be 4.75% (95% CI 3.05%; 6.75%). We considered this to be an appropriate source for VE waning against infection because there is minimal to no protection against COVID-19 infection a year after receiving vaccination, which implies that waning of annual vaccination VE would be similar to the waning of VE of those who received initial doses of COVID-19 vaccine. However, as protection against hospitalization is longer lasting, a study by Andersson et al., (2024)^22^ which examined the effectiveness of XBB.1.5 containing COVID-19 mRNA vaccines was used to estimate the base case rate of waning of VE against hospitalization. This study found that VE against hospitalization waned at a rate of 1.7% over a three-week period, which is equivalent to 2.46% per month. In sensitivity analyses, the estimate from Higdon et al, against hospitalization (1.37%), as well as an alternate source which examine the durability of the mRNA-1273 XBB.1.5 vaccine (3.87%)^26^ were tested.

### Probabilities for the COVID-19 Consequences Tree

Key probability inputs are summarized in Table 4 and the text. Further details are provided in the Technical Appendix.

**Table 4.**
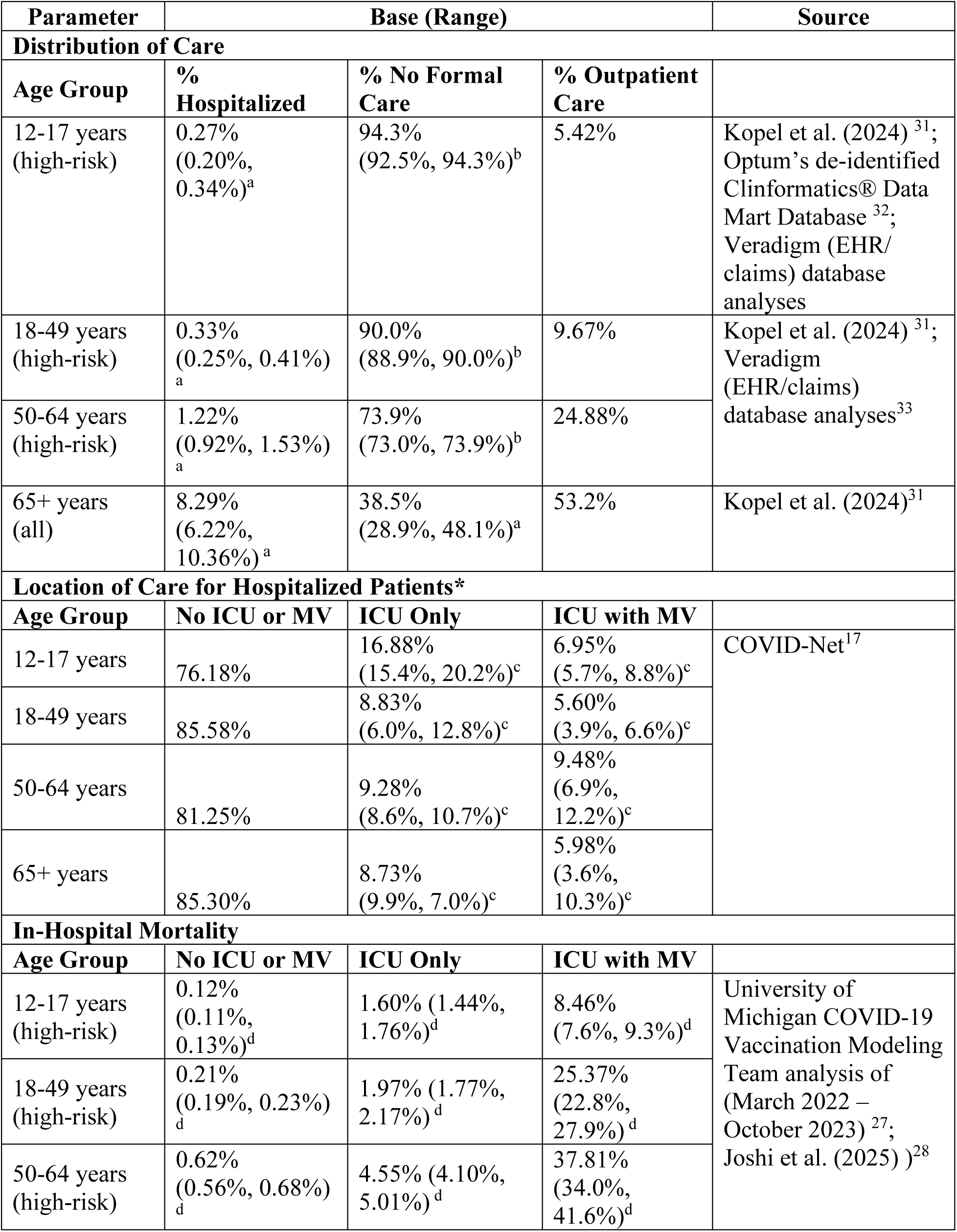
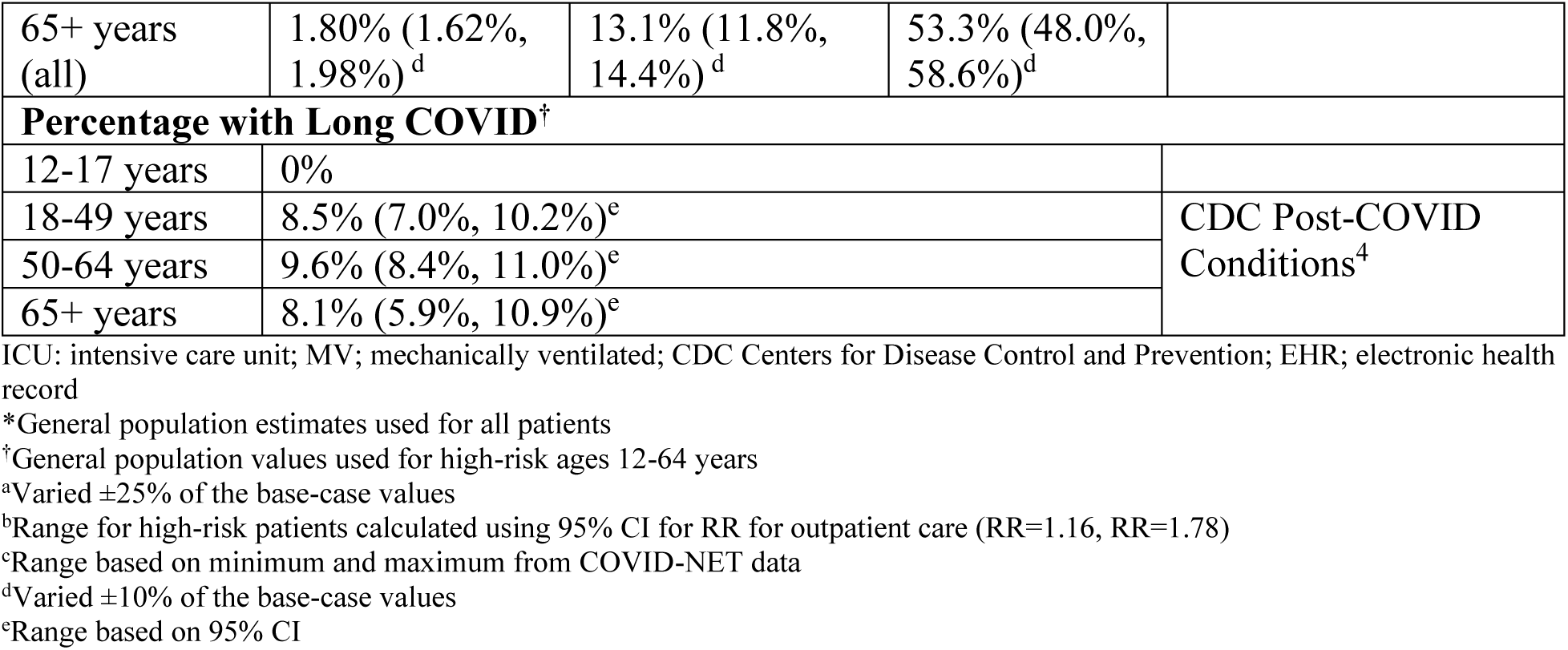
Key decision tree probabilities, base case and range for deterministic sensitivity analyses.

Patients with symptomatic COVID-19 infections are divided by their highest level of medical care received: 1) No formal health care (not hospitalized nor was outpatient care received); 2) Outpatient care (not hospitalized but did receive outpatient physician visits or emergency department visits without hospital admission); and 3) Inpatient care. These age-specific probabilities were estimated based on database analyses and adjusted to represent the high-risk population for ages 12 to 64 years as described in the Technical Appendix. Hospitalized patients are further stratified by location of care, including general ward (no intensive care unit [ICU] or ventilation required), ICU (excluding extracorpeal membrane oxygenation or invasive ventilation), or ICU with mechanical ventilation based on data from the COVID-Net surveillance system.^17^ In absence of data, the proportions for the high-risk populations were assumed to be equivalent to the general population.

Hospitalized patients may die, be discharged and then readmitted, or simply discharged. Estimates of in-hospital mortality for the general population^27^, stratified by age and in-hospital location of care, were based on COVID-NET data from March 2022 – October 2023 as analyzed by the University of Michigan COVID-19 Vaccination Modeling Team.^27^ Based on Joshi et al. (2025) ^28^, it was conservatively assumed that the relative risk for in-hospital mortality for high-risk patients relative to non-high-risk patients was 1.23, considering an estimate derived for patients with diabetes. The COVID-NET surveillance system tracks only in-hospital deaths and not post-discharge mortality. The 30-day readmission and post-discharge mortality probabilities were estimated from a meta-analysis for the general population.^29^ The readmission rate for the high-risk populations was calculated using the RR for patients with diabetes (RR=1.27) from Joshi et al. (2025)^28^ and a weighted^16^ average of high-risk and general population of 9.35% (range: 4.1%, 9.28%) was used. A post-discharge mortality of 7.87% (95% CI: 2.8% to 13.0%) was assumed to be the same for both the high-risk and general populations and applied to all age groups.

Data from the CDC based on the US Census Bureau Household Pulse Survey^4^ were used to estimate the prevalence of long COVID for patients receiving either outpatient or hospital-based care. Estimates are based on the average percentages of adults who report currently experiencing long COVID among those who ever had COVID based on data from August 20-September 16, 2024^30^. As data were limited to adults, children were conservatively assumed to not experience long COVID. General population estimates are applied to high-risk populations.

The probability of infection-related myocarditis is described in the Technical Appendix.

### Sensitivity Analyses

Deterministic sensitivity analyses were performed to assess the impact of key variables on estimated symptomatic infections, COVID-19 hospitalizations, and deaths. The monthly incidence of COVID-19 infection was increased and decreased by 25% in approximation of uncertainty in COVID-19 incidence as full annual 2024/2025 surveillance data were not available at time of the conduct of the study. The ranges used for the probability of infection consequences are displayed in Table 3. Vaccine coverage was also varied. Upper bounds were estimated by using the ratio between 2023-2024 coverage data for high-risk individuals and the general population obtained from COVIDVaxView^34^ and applying this ratio to the base-case coverage rates (Technical Appendix). The lower bounds of vaccine coverage were estimated using data from the UK; during the Spring 2022 booster campaign in the UK, when vaccines were offered to all adults ≥75 years and immunocompromised individuals, coverage was approximately 79%^35^. Following the policy shift to targeting high-risk groups only, only 53% of those ≥75 years^36^ received the spring booster in the spring of 2023, yielding a ratio of 0.668, which was applied to the base-case vaccine coverage rates (Technical Appendix). Additional vaccination coverage rate scenarios were tested where different magnitudes of increases of vaccine uptake were explored (Technical Appendix). These increases were however less than the upper bound scenario, thus results of these are only described in the Technical Appendix. Finally, the initial VE inputs and the monthly waning rates were varied as described above. Several scenario analyses described below were conducted (further detail on model inputs are presented in the Technical appendix). An interim 2024/2025 incidence scenario was created based on COVID-NET hospitalizations between September 2024 and May 2025. Two scenario analyses were conducted for VE: 1) rVE against hospitalization for mRNA-1283 versus mRNA-1273 were set to be equal to the rVE against infections; and 2) VE against symptomatic COVID-19 infection for adults ages 18 years and older from ICATT study.^37^ The model cannot estimate any benefit associated with changes in transmission, which may include the reduction in secondary transmission when a vaccinated person develops an infection. This effect was demonstrated by the CDC RIGHT study^21^, which estimated that if infected people had been vaccinated within the prior 6 months, their risk of transmitting to household contacts was reduced compared to the who had not been vaccinated (adjusted RR 0.55; VE: 45%). These results were therefore used to create an indirect benefit scenario. Several scenario analyses were also performed specific to high-risk individuals, including assuming that 50% of 12-17 year-olds are considered high-risk, that the likelihood of receiving outpatient care is equivalent to the general population, that the RR of inpatient mortality is 1.62 based on cardiovascular disease^28^, and using the percentage hospitalized based on the RR derived from Wilson et al. (2025) (RR=1.48).

For the analyses comparing mRNA-1283 to mRNA-1273 and BNT-162b2, a range of rVE values were used to calculate the ranges used for sensitivity analyses as shown in Table 3.

## Results

### Base case analysis

As shown in Table 5, use of an annual dose of mRNA-1283 is estimated to reduce the number of symptomatic infections, outpatient episodes, hospitalizations, deaths, and cases of long COVID in the US population by 2.86 million, 825,628, 171,208, 22,353, and 81,570 compared to no vaccine, respectively. The corresponding numbers needed to vaccinate (NNV) to avert one COVID-19 symptomatic infection, outpatient episode, hospitalization, death and long COVID were estimated at 19, 65, 314, 2,411, and 661 respectively. The majority of severe COVID-19 cases, defined as hospitalizations and deaths, were prevented among adults aged 65 years and older, accounting for 152,297 hospitalizations and 20,212 deaths. These figures represent 90.4% of all hospitalizations and 89.0% of all deaths, respectively. Second dose coverage has been low, but use of semi-annual dosing in those 65 years and older is estimated to increase the number averted to 2.91 million symptomatic infections, 173,544 hospitalizations, and 22,663 deaths in the overall target population of 12 to 64 high-risk and 65 years and older.

**Table 5:** Health outcomes and health outcomes averted for the base case analysis with annual vaccination and for the scenario with semi-annual vaccination for ages 65 years and older.

| Strategy | Health outcomes |  |  |  |  | Health outcomes averted |  |  |  |  |
| --- | --- | --- | --- | --- | --- | --- | --- | --- | --- | --- |
|  | Cases | Outpatient | Long COVID | Hosp. | Deaths | Cases | Outpatient | Long Covid | Hosp. | Deaths |
| No vaccine | 38,519,798 | 8,228,024 | 760,310 | 791,068 | 100,874 | - | - | - | - | - |
| mRNA-1283 COVID-19 vaccine, 1- dose | 35,664,235 | 7,402,396 | 678,740 | 619,860 | 78,521 | 2,855,564 | 825,628 | 81,570 | 171,208 | 22,353 |
| No vaccine | 38,519,798 | 8,228,024 | 760,310 | 791,068 | 100,874 | - | - | - | - | - |
| mRNA-1283 COVID-19 vaccine, 2- dose | 35,609,940 | 7,371,342 | 676,072 | 617,524 | 78,211 | 2,909,858 | 856,682 | 84,239 | 173,544 | 22,663 |

Hosp.: Hospitalizations; 1-dose: annual; 2-dose: annual for <65; semi-annual for ages 65+

### Deterministic Sensitivity Analyses

The impact of deterministic sensitivity analyses on the number of symptomatic infections, hospitalizations, and deaths prevented is summarized in the tornado diagram in Figure 3. Details are provided in the Technical Appendix. Overall, variation in vaccine coverage has the greatest impact on the estimated number of all outcomes prevented. Varying the percent of people with infections who are hospitalized has the next largest impact on the number of hospitalizations and deaths averted. Varying the monthly infection incidence rates by 25% has the third largest impact on hospitalizations and deaths averted and the second largest impact on infections averted. Next, initial hospitalization VE and waning rate of hospitalization VE impacts the number of hospitalizations and deaths averted. Only the number of infections prevented is impacted by variation in the initial infection VE and the waning rate of infection VE as the VE against hospitalization remains the same.

**Figure 3:**
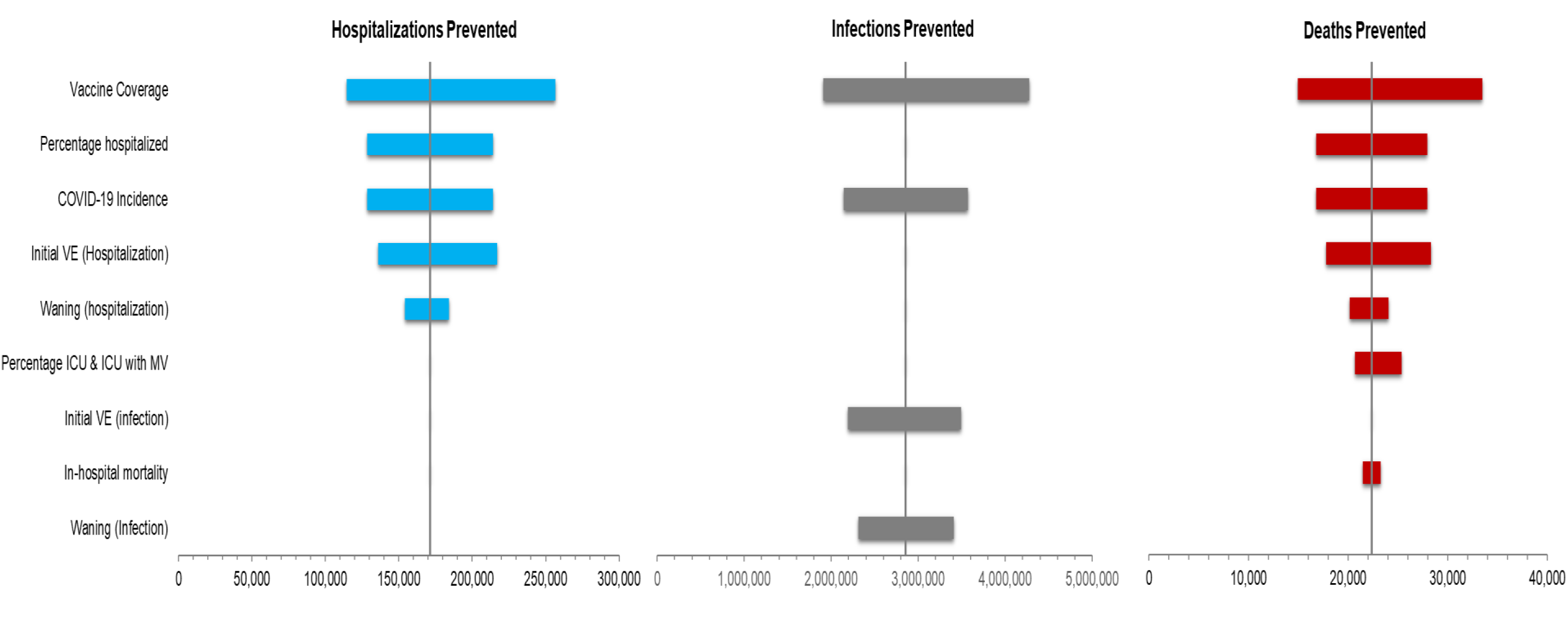
Tornados: Impact of deterministic sensitivity analyses on the number of hospitalizations, infections and deaths prevented.

### Scenario Analyses

The results of the scenario analyses are shown in the Technical Appendix. Changing the incidence of infection to the interim 2024/25 scenario had the largest impact on results as it decreased the averted infections, hospitalizations, and deaths by 53%, 55% and 55% respectively. On the other hand, accounting for the effect of reduction of transmission (i.e. indirect benefit) increased the number of infections, hospitalizations and deaths prevented by 45%, 29% and 29% respectively. Setting the rVE of hospitalization to be the same as the rVE of infection decreases the number of hospitalizations and deaths averted by 17%. The number of symptomatic infections averted increased by 14% when the VE against infections from the ICATT study is used in place of base case values as this increased the estimate of initial VE against infection for mRNA-1283. All remaining scenario analyses had minimal impact on the predicted clinical impact of the vaccine.

### Comparator Scenario Analyses

With annual dosing, the next generation mRNA-1283 is estimated to prevent 0.56 million more symptomatic infections, 45,500 more hospitalizations, and 5,921 more deaths than the original mRNA-1273 vaccine. The additional hospitalizations prevented by mRNA-1283 over mRNA-1273 correspond to an incremental NNV of 114, which means that 114 more persons would need to be vaccinated with mRNA-1273 to prevent one hospitalization when compared with mRNA-1283. 39,483 or 87% of these additional prevented hospitalizations were prevented among adults 65 years and older. The uncertainty is reflected in the ranges in Table 6. along with an analyses including semi-annual dosing for those 65 years and older. Furthermore, mRNA-1283 is expected to prevent 0.79 million more symptomatic infections, 58,073 more hospitalizations and 7,565 deaths than BNT162b2. Again, when considering additional hospitalizations prevented by mRNA-1283 over BNT162b2, the corresponding incremental NNV was estimated at 162, which means that 162 more persons would need to be vaccinated with BNT162b2 to prevent a hospitalization when compared with mRNA-1283. 50,766 or 87% of these additional prevented hospitalizations were prevented among adults 65 years and older. The predicted range for this comparison and the comparison with semi-annual dosing for those 65 years and over are displayed in Table 7.

**Table 6:** Health outcomes and health outcomes averted for the mRNA-1283 compared to mRNA-1273.

| Strategy | Health outcomes |  |  |  |  | Health outcomes averted |  |  |  |  |
| --- | --- | --- | --- | --- | --- | --- | --- | --- | --- | --- |
|  | Cases | Outpatient | Long COVID | Hosp. | Deaths | Cases | Outpatient | Long Covid | Hosp. | Deaths |
| **mRNA-1273 COVID-19 vaccine, 1-dose** | 36,224,322 | 7,531,194 | 693,007 | 665,360 | 84,443 | - | - | - | - | - |
| **mRNA-1283 COVID-19 vaccine, 1 dose (Base-Case VE)** | 35,664,235 | 7,402,396 | 678,740 | 619,860 | 78,521 | 560,088 | 128,798 | 14,267 | 45,500 | 5,921 |
| **mRNA-1283 COVID-19 vaccine, 1 dose (95% CI LB VE)** | 36,183,191 | 7,524,383 | 692,383 | 665,359 | 84,442 | 41,131 | 6,811 | 625 | 2 | 0 |
| **mRNA-1283 COVID-19 vaccine, 1 dose (95% CI UB VE)** | 35,143,315 | 7,245,742 | 663,382 | 588,707 | 74,467 | 1,081,007 | 285,452 | 29,625 | 76,653 | 9,976 |
| **mRNA-1273, 2-dose** | 36,170,029 | 7,500,142 | 690,339 | 663,024 | 84,132 | - | - | - | - | - |
| **mRNA-1283 COVID-19 vaccine, 2- dose** | 35,609,940 | 7,371,342 | 676,072 | 617,524 | 78,211 | 560,089 | 128,799 | 14,268 | 45,500 | 5,921 |

Hosp.: Hospitalizations; 1-dose: annual; 2-dose: annual for <65; semi-annual for ages 65+; CI: Confidence interval; LB: Lower bound; VE: Vaccine effectiveness; UB: Upper bound

**Table 7:** Health outcomes and health outcomes averted for the mRNA-1283 compared to BNT-126b2.

| Strategy | Health outcomes |  |  |  |  | Health outcomes averted* |  |  |  |  |
| --- | --- | --- | --- | --- | --- | --- | --- | --- | --- | --- |
|  | Cases | Outpatient | Long COVID | Hosp. | Deaths | Cases | Outpatient | Long Covid | Hosp. | Deaths |
| **mRNA-1283 COVID-19 vaccine, 1-dose** | 35,664,235 | 7,402,396 | 678,740 | 619,860 | 78,521 | - | - | - | - | - |
| **BNT162b2 (Base-Case VE)** | 36,452,116 | 7,626,590 | 701,721 | 677,933 | 86,086 | 787,882 | 224,195 | 22,981 | 58,073 | 7,565 |
| **BNT162b2 (95% CI LB VE)** | 37,126,466 | 7,838,515 | 722,463 | 720,121 | 91,628 | 1,462,232 | 436,119 | 43,723 | 100,261 | 13,107 |
| **BNT162b2 (95% CI UB VE)** | 35,813,257 | 7,444,568 | 682,478 | 622,321 | 78,840 | 149,023 | 42,172 | 3,738 | 2,461 | 319 |
| **mRNA-1283 COVID-19 vaccine, 2- dose** | 35,609,940 | 7,371,342 | 676,072 | 617,524 | 78,211 |  |  |  |  |  |
| **BNT162b2, 2-dose** | 36,398,591 | 7,596,010 | 699,091 | 675,597 | 85,776 | 788,651 | 224,668 | 23,019 | 58,073 | 7,565 |

Hosp.: Hospitalizations; 1-dose: annual; 2-dose: annual for <65; semi-annual for ages 65+; CI: Confidence interval; LB: Lower bound; VE: Vaccine effectiveness; UB: Upper bound
\*Additionally prevented health outcomes of by mRNA-1283 compared to BNT162b2

## Discussion

During the 2024/2025 season in the United States, we estimate in this interim analysis that the next-generation Moderna COVID-19 mRNA-1283 vaccine would have prevented, in its licensed target population (adults aged ≥65 years and individuals aged 12–64 years with underlying medical conditions), approximately 2.9 (1.3 – 4.3) million symptomatic COVID-19 infections, 170,000 (77,000 - 260,000) hospitalizations, and 22,350 (10,050 - 33,480) deaths compared to no vaccination.

Compared with the originally approved mRNA vaccines (mRNA-1273 and BNT162b2), we estimate that mRNA-1283 would have prevented a substantial additional number of symptomatic infections, hospitalizations, and deaths. Specifically, compared to BNT162b2, we estimate that mRNA-1283 would have averted an additional 0.79 million symptomatic infections, 58,000 hospitalizations, and 7,565 deaths. Compared to mRNA-1273, we estimate that mRNA-1283 would have averted an additional 0.56 million symptomatic infections, 46,000 hospitalizations, and 5,921 deaths.

These interim estimates are consistent with findings from a recent Canadian public health impact analysis showing a large reduction in COVID-19 infections and severe COVID-19 cases with mRNA-1283when compared to no vaccination and substantial additional COVID-19 events prevented when compared to current COVID-19 mRNA vaccines.^38^.

Combined with multiple scenario and sensitivity analyses confirming the robustness of the current model, the results suggest that mRNA-1283 represents a valuable option to optimize US COVID-19 vaccination strategies—particularly for older adults (≥65 years) and individuals with high-risk comorbidities who remain most vulnerable to severe disease. Furthermore, the differences in COVID-19 outcomes prevented by mRNA-1283 compared to each mRNA-1273 and BNT162b2 underscore that the choice of mRNA COVID-19 vaccine likely result in substantial differences in clinical impact.

This interim analysis has several limitations. First, the real-world effectiveness and safety of mRNA-1283 have not yet been established. The VE input was based on interim vaccine effectiveness estimates for mRNA-1273 targeting the KP.2 variant, combined with the rVE between mRNA-1283 and mRNA-1273. The rVE estimate for hospitalization was derived from a post-hoc analysis of FDA-defined severe COVID-19, which only included a total of 55 FDA-defined severe COVID-19 events with the majority of cases due to blood pressure and oxygen saturation abnormalities. Additionally, initial rVE estimates for mRNA-1283 versus BNT162b2 were based on indirect treatment comparisons^12^ and should be validated and refined with real-world head-to-head data.

The model assumed equal waning across all three vaccines, which may lead to conservative estimates for mRNA-1283, particularly in older adults, given that the Phase 3 trial findings show higher immunogenicity for mRNA-1283 at day 181 compared to mRNA-1273 at day 91^39^.

For model calibration, 2023/2024 season CDC-reported hospitalization rates were used as the base case due to full 2024/2025 season data not being available at the time of conducting this study and considering the observed multi-peak pattern and sustained incidence between peaks during the past 2023/2024 season. However, substantial uncertainty remains regarding future COVID-19 incidence. A future update of this analysis may consider full 2024/2025 incidence data or average rates across multiple post-pandemic seasons.

This interim assessment suggests that the next-generation COVID-19 mRNA-1283 vaccine could substantially reduce the clinical burden of COVID-19 among high-risk populations in the US—specifically, individuals aged 12–64 years with underlying conditions and all adults aged ≥65 years. These benefits were observed both in comparison to no vaccination and to the originally approved mRNA vaccines. As such, mRNA-1283 may represent a valuable option for optimizing COVID-19 vaccination policy in the US for those at highest risk of severe outcomes.

### Transparency

- **Declaration of Funding:** This study was supported by Moderna, Inc.
- **Declaration of competing interests:** MK is a shareholder in Quadrant Health Economics Inc, which was contracted by Moderna, Inc. to conduct this study. AL, KF, MW and SC are consultants to Quadrant Health Economics Inc. KJ, NV, EB and MB are employed by Moderna, Inc. and may hold stock/stock options in the company.

## Data Availability

All data produced in the present work are contained in the manuscript.

