## Supplemental File for "Modeling the Anticipated Public Health Benefits of the Next-Generation COVID-19 mRNA-1283 vaccine: An Interim U.S. Population-Level Impact Assessment"

- Technical Appendix –

### 1 Estimation of Incidence of Infection (No Vaccination)

As the hospitalization data from the CDC reflects a partially vaccinated population, information on vaccination coverage, VE, and the probability of hospitalization must be used as well as the target hospitalization rates. The process is described below.

**Step 1**: Develop the age-specific targets for the monthly rate of hospitalizations by age group for a one-year period. As hospitalization rates for one full year, i.e. 12 months, are needed, for the base case, the hospitalization rates for September 2023 to August 2024 from COVID-NET^[1]^ were used.^1^ The rates per 100,000 are displayed in Table 1 below.

Table 1. Target hospitalization rates per 100,000 for the calculation of infection incidence in the model^1^

| **Age Group (Years)** | **Sep-23** | **Oct-23** | **Nov-23** | **Dec-23** | **Jan-24** | **Feb-24** | **Mar-24** | **Apr-24** | **May-24** | **Jun-24** | **Jul-24** | **Aug-24** |
| --- | --- | --- | --- | --- | --- | --- | --- | --- | --- | --- | --- | --- |
| 12-17 | 1.8 | 0.8 | 1.6 | 2.5 | 2.8 | 2.7 | 1.4 | 0.3 | 0.6 | 0.4 | 1.2 | 1.7 |
| 18-49 | 5.5 | 5.2 | 6.0 | 8.2 | 9.0 | 5.8 | 3.0 | 1.6 | 1.6 | 2.6 | 5.3 | 6.7 |
| 50-64 | 15.7 | 15.1 | 18.7 | 23.8 | 26.3 | 16.1 | 8.8 | 5.0 | 4.4 | 6.4 | 13.1 | 17.0 |
| ≥ 65 | 80.5 | 80.6 | 90.7 | 122.3 | 112.7 | 68.7 | 42.9 | 25.6 | 24.3 | 39.1 | 66.4 | 82.3 |

**Step 2**: Enter the age-specific probability of hospitalization and the related proportion of symptomatic cases that are seeking care. The same probabilities are used for the cost-effectiveness model of the symptomatic infection in the no vaccination arm and the incidence calculation.

**Step 3**: Enter the assumed vaccination coverage.

The monthly vaccine coverage rate for September 2023 to August 2024, displayed in Table 2, was estimated from the vaccine coverage of the XBB.1.5 vaccines from the CDC VaxView database.^2^ Data were available from the last week of September 2023, when XBB.1.5 vaccine became available, up to the week ending May 11, 2024, and were disaggregated by age groups. Administration of the first dose was assumed to begin September 3, 2023 and end May 11, 2024. In the spring of 2024, individuals age 65 years and over were eligible to receive a second dose of the COVID-19 vaccines. Data on coverage of a second dose of the XBB.1.5 vaccine was also available for individuals aged 65 years and older, and began April 1, 2024 and end July 27, 2024. Figure 1 and Figure 2 provides detailed vaccine coverage over time for the first and second doses, respectively.

A small proportion of people aged 65 years and older in the US were vaccinated with a second dose between April and July 2024 (See Figure 2). The model only accommodates one dose for the calculations of infection incidence, so the single dose coverage rates were increased to account for second doses in spring 2024. Overall, excluding these vaccinations or including them made a small difference to the final infection incidence rates.

Figure 1. First dose COVID-19 vaccine coverage during season 2023-2024, by age groups

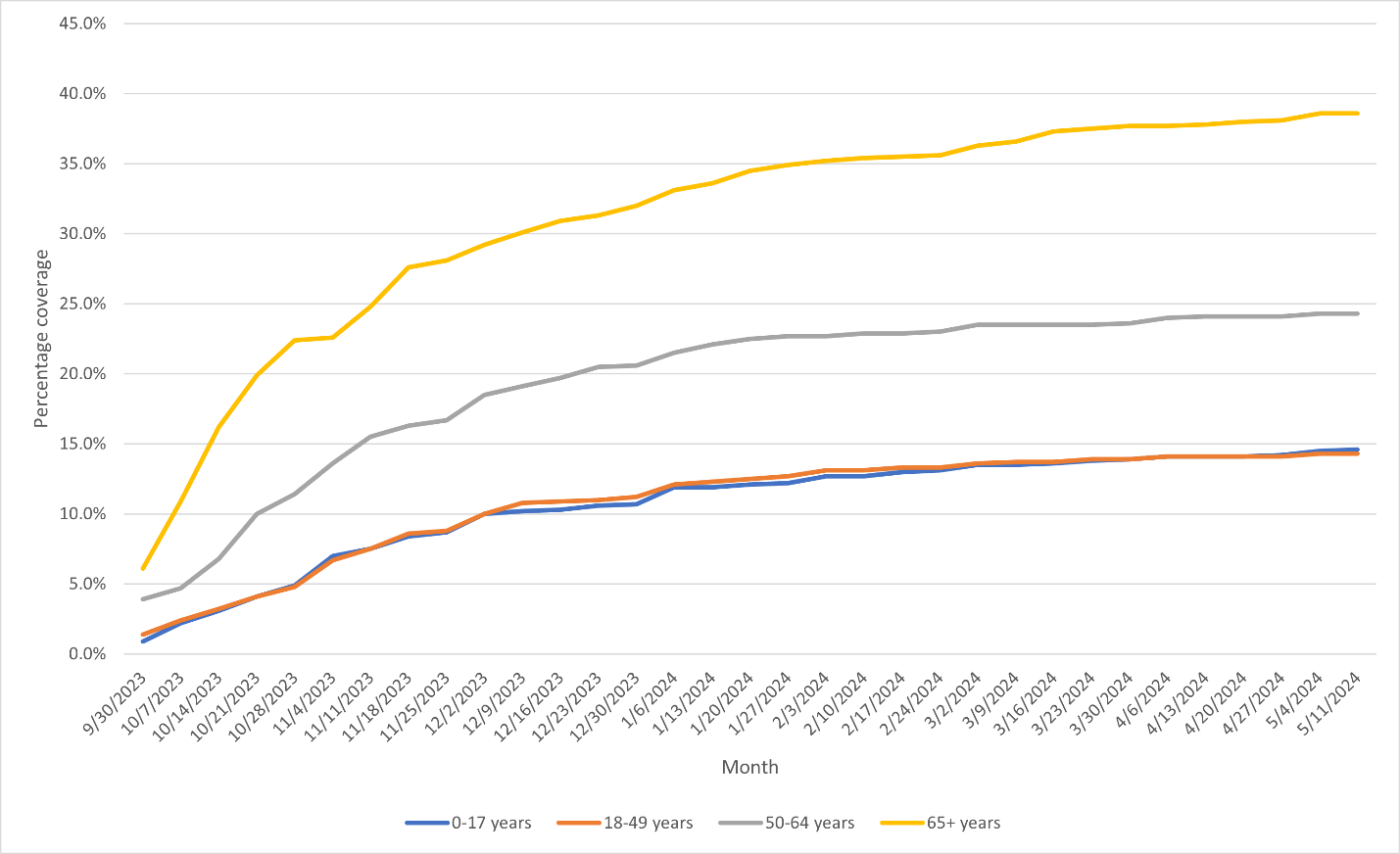

Figure 2. Second dose, COVID-19 vaccine coverage during season 2023-2024

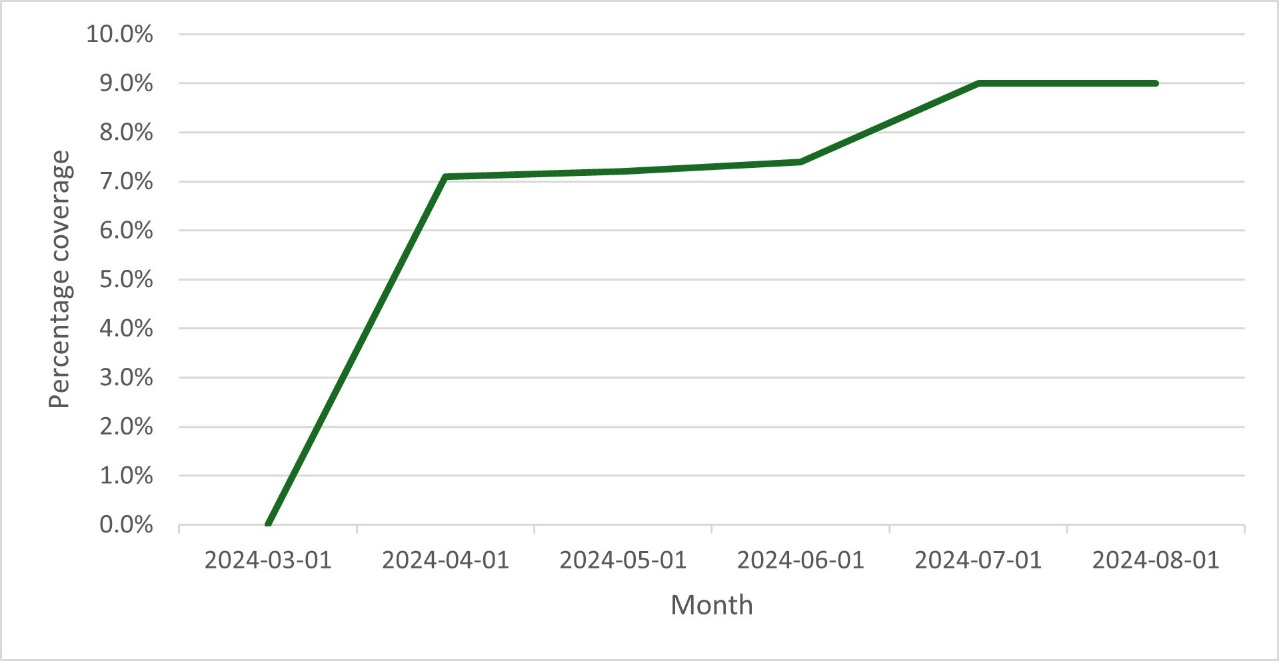

Table 2. Monthly coverage rates used in the calculation of infection incidence in the static model

| **Date** | **12-17 years** | **18-49 years** | **50-64 years** | **65+ years** |
| --- | --- | --- | --- | --- |
| 30-Sep-2023 | 0.9% | 1.4% | 3.9% | 6.1% |
| 31-Oct-2023 | 7.6% | 5.6% | 12.3% | 22.5% |
| 30-Nov-2023 | 12.3% | 9.7% | 18.0% | 28.9% |
| 31-Dec-2023 | 14.2% | 11.3% | 20.7% | 32.2% |
| 31-Jan-2024 | 15.7% | 12.9% | 22.7% | 35.1% |
| 29-Feb-2024 | 16.6% | 13.5% | 23.4% | 36.1% |
| 31-Mar-2024 | 17.2% | 13.9% | 23.7% | 37.7% |
| 30-Apr-2024 | 17.6% | 14.2% | 24.2% | 45.4% |
| 31-May-2024 | 18.1% | 14.3% | 24.3% | 45.8% |
| 30-Jun-2024 | 18.1% | 14.3% | 24.3% | 46.0% |
| 31-Jul-2024 | 18.1% | 14.3% | 24.3% | 47.6% |
| 31-Aug-2024 | 18.1% | 14.3% | 24.3% | 47.6% |

**Step 4**: Enter the assumed initial vaccine effectiveness (VE) against infection and hospitalization and the monthly linear waning rate over time. **See Section 2.2**.

In the model calculation sheets, an initial set of monthly incidence rates of symptomatic infection without seasonal vaccination is used to calculate the number of hospitalizations by month and age group. These estimated hospitalization counts are compared to the target number of hospitalizations by month and age group and the ratio of target to model estimated hospitalizations is calculated for each month and age group. These ratios are multiplied by the initial set of monthly incidence rates to determine the calibrated monthly incidence rates by age group that are required to estimate the target number of hospitalizations, given the other model inputs are held constant. These final incidence rates are then output and can be copied directly into the static CEA model. The base case incidence values are displayed in Table 3.

Table 3. Base case incidence of symptomatic infection (no vaccination arm) for the static model (% infected)

| **Age group (years)** | **Sep-23** | **Oct-23** | **Nov-23** | **Dec-23** | **Jan-24** | **Feb-24** | **Mar-24** | **Apr-24** | **May-24** | **Jun-24** | **Jul-24** | **Aug-24** |
| --- | --- | --- | --- | --- | --- | --- | --- | --- | --- | --- | --- | --- |
| 12-17 | 0.98 | 0.46 | 0.95 | 1.49 | 1.69 | 1.63 | 0.85 | 0.18 | 0.36 | 0.24 | 0.72 | 1.01 |
| 18-49 | 1.96 | 1.90 | 2.25 | 3.10 | 3.43 | 2.21 | 1.14 | 0.61 | 0.61 | 0.98 | 1.99 | 2.51 |
| 50-64 | 1.43 | 1.45 | 1.86 | 2.40 | 2.68 | 1.63 | 0.89 | 0.50 | 0.44 | 0.64 | 1.30 | 1.67 |
| 65+ | 1.01 | 1.13 | 1.32 | 1.82 | 1.69 | 1.03 | 0.64 | 0.40 | 0.38 | 0.60 | 1.02 | 1.25 |

### 2 Vaccine Effectiveness

#### 2.1 Calculation of Vaccine Effectiveness with the Model

Vaccine effectiveness declines linearly on a monthly basis within the model. A portion of the cohort can receive a new vaccine in each month of the time horizon. In order to incorporate waning of effectiveness, the effectiveness calculation is a function of the fraction of the age group vaccinated each month and vaccine effectiveness. Vaccine effectiveness is the sum of the following:

$Effectiveness={Cov}_{Cur}\times{Eff}_{Cur}+{Cov}_{Cur-1}\times{Eff}_{Cur-1}+ {Cov}_{Cur-2}\times{Eff}_{Cur-2}+\ldots+ {Cov}_{Cur-9}\times{Eff}_{Cur-9}$

Where:

Cov_cur_ = fraction of age group vaccinated in the current month

Cov_cur1_ = fraction of age group vaccinated in the month before the current month

Cov_cur2_ = fraction of the age group vaccinated two months before the current month

Eff_cur_ = effectiveness in the current month

Eff_cur1_ = effectiveness one month after initial vaccination

Eff_cur2_ = effectiveness two months after initial vaccination

#### 2.2 Inputs for the Estimation of Incidence

The 2023-2024 COVID-19 VE against infection and hospitalization for adults (aged 18 years and over) were based on a market mix of the COVID-19 mRNA vaccines mRNA-1273 and BNT162b2 administered during 2023-2024. Both VE outcomes of the Moderna XBB.1.5 vaccine were estimated based on a study by Kopel et al., (2024)^3^ that estimated the VE in those ≥18 years old receiving the vaccine between September 13, 2023 and December 15, 2023 relative to individuals that did not receive the XBB.1.5 vaccine during the same time period, independent of prior vaccination history. As the median follow-up time was 70 days post-vaccination, values for both outcomes were scaled back by 56 days to 14 days post vaccination to approximate the maximum level of protection as described in the sections below.

*Infection:*

The 2023-2024 VE for the Moderna XBB.1.5 vaccine against medically-attended SARS-CoV-2 infection at 70 days (33.1%)^3^ was used as a proxy for VE against COVID-19 symptomatic infection as this outcome was not directly measured in the Kopel et al study. As Higdon et al., (2022)^4^ approximated the aVE against infection to wane at a rate of 4.75% (95% CI 3.05%; 6.75%) monthly, the monthly waning rate of 4.75% was applied to 56 days and added to the 2023-2024 VE for the Moderna vaccine from Kopel to approximate the VE against infection at 2 weeks post-administration (41.8%). These values were used for adults aged 18 years and over.

The 2023-2024 VE for the Pfizer XBB.1.5 vaccine were estimated based on adjusted rVE values between Moderna and Pfizer vaccine values. At time of calculation, values were available for bivalent original and Omicron BA.4/5 containing mRNA-1273.222 versus BNT1162b2 , and were therefore used. rVE for outpatient visits were used as a proxy for symptomatic infection (rVE=5.1%, 95% CI 3.2-6.9%).^5^ This resulted in a 2023-2024 Pfizer XBB.1.5 VE value against infection of 38.7% in those aged 18 years and over.

The Moderna and Pfizer 2023-2024 COVID-19 VE values were then weighed by the estimated market shares of each (48% and 52%, respectively),^6^ to obtain a weighted 2023-2024 COVID-19 VE for the vaccines (40.2%).

For the age group 12 to 17 years, the 2023-2024 COVID-19 VE value was obtained from a study by Link-Gelles.^7^ The 2023-2024 COVID-19 VE for the age group of 5-17 years was 71.0% and assumed for those 12-17 years in the model. As VE was measured between 7-59 days post XBB.1.5 vaccine administration, adjustments to 2 weeks post-administration was not required.

*Hospitalization:*

The monthly waning rate of 2.46% calculated from Andersson et al., (2024)^8^ was combined with the 2023-2024 Moderna XBB.1.5 VE value against hospitalization from Kopel et al., (2024),^3^ to approximate the 2023-2024 Moderna XBB.1.5 VE against hospitalization at 14 days post-administration (64.7%).

Similar to the 2023-2024 Pfizer XBB.1.5 VE against infection, the 2023-2024 Pfizer XBB.1.5 VE against hospitalization was estimated by applying the hospitalization rVE value between the bivalent versions of mRNA-1273.222 versus BNT162b2 as a proxy (rVE=9.8%, 95% CI 2.6-16.4%).^5^ This resulted in a 2023-2024 Pfizer XBB.1.5 initial VE value against hospitalization of 60.9%.

The weighted 2023-2024 COVID-19 VE value against hospitalization using the market share values presented above is therefore 62.7%.

Table 4 summarizes the market mix VE used for calculation of the incidence of symptomatic infection without vaccination.

Table 4. Market mix VE used in the calculation of infection incidence in the model

| **Age Group** | **Infection** | | **Hospitalization** | |
| --- | --- | --- | --- | --- |
|  | **Initial VE** | **Waning** | **Initial VE** | **Waning** |
| 12 to 17 years | 0.7100 | 0.0475 | 0.7100 | 0.0246 |
| 18-49 years | 0.4020 | 0.0475 | 0.6270 | 0.0246 |
| 50-64 years | 0.4020 | 0.0475 | 0.6270 | 0.0246 |
| ≥65 years | 0.4020 | 0.0475 | 0.6270 | 0.0246 |

VE: Vaccine effectiveness

#### 2.3 Calculation of Incremental VE against hospitalization

The model assumes the VE against infection to be lower than the VE against hospitalization and thus applies an incremental VE against hospitalization for infections occurring in vaccinated in the vaccination arm. The hospitalization VE values are adjusted in the model to reflect the incremental protection against hospitalization above the protection against infection. In other words, VE values were adjusted to account for cases of hospitalization that are prevented due to the decrease in infections with vaccinations. This was to ensure that protection against hospitalizations is not double counted. The equations used for this are presented below.

For each age group, we define the following vaccine effectiveness variables and relationship between the variables. The superscripts and subscripts for age group are removed for clarity.

**Definitions**

${VE}_{1}$ = Vaccine effectiveness against infection

${VE}_{2}$ = ‘Total’ Vaccine effectiveness against hospitalization

${VE}_{2}^{*}$= ‘Additional’ Vaccine effectiveness against hospitalization

We assume ${VE}_{2}^{*}=0$ if there is no additional benefit against hospitalization

**Define**

$$\left[ 1-{VE}_{2} \right]= \left[ 1-{VE}_{1} \right]\times\left[ 1-{VE}_{2}^{*} \right]$$

Isolate and solve for ${VE}_{2}^{*}$

$$\left[ 1-{VE}_{2}^{*} \right]=\frac{\left[ 1-{VE}_{2} \right]}{\left[ 1-{VE}_{1} \right]}$$

$${VE}_{2}^{*}= 1- \frac{\left[ 1-{VE}_{2} \right]}{\left[ 1-{VE}_{1} \right]}$$

### 3 Vaccine Coverage

A figure showing the vaccine uptake by month used for the analytic time horizon, based on data from September 2024 to February 2025 from COVIDVaxView,^9,10^ is displayed below. Vaccination uptake by month for the upper and lower bound vaccination coverage rate scenarios tested in deterministic sensitivity analysis are shown in Table 5. The uptake of the second dose for the scenario where semi-annual vaccination of those 65 years and older is tested is shown in Section 1 (Figure 2).

Based on observed increases of CDC reported COVID-19 vaccination coverage rates in persons 65 years and older between season 2023/2024 and 2024/2025, two additional scenarios for 1-dose vaccination strategies were studied assuming an increase in vaccination coverage rates by 10% in the overall population, i.e. 12 to 64 years high-risk and 65 years and older, as well as 65 years and older only. The corresponding vaccination uptake is shown in Table 6.

Figure 3. First seasonal dose vaccine coverage by age used for the analytic time horizon.

Percent Coverage

|  | 12-17 years | 18-49 years | 50-64 years | 65+ years |
| --- | --- | --- | --- | --- |
| 2024-09-01 | 0% | 0% | 0% | 0% |
| 2024-09-02 | 0.141176% | 0.15% | 0.333333% | 0.683333% |
| 2024-09-03 | 0.282353% | 0.3% | 0.666667% | 1.36667% |
| 2024-09-04 | 0.423529% | 0.45% | 1% | 2.05% |
| 2024-09-05 | 0.564706% | 0.6% | 1.33333% | 2.73333% |
| 2024-09-06 | 0.705882% | 0.75% | 1.66667% | 3.41667% |
| 2024-09-07 | 0.847059% | 0.9% | 2% | 4.1% |
| 2024-09-08 | 0.988235% | 1% | 2.15714% | 4.62857% |
| 2024-09-09 | 1.12941% | 1.1% | 2.31429% | 5.15714% |
| 2024-09-10 | 1.27059% | 1.2% | 2.47143% | 5.68571% |
| 2024-09-11 | 1.41176% | 1.3% | 2.62857% | 6.21429% |
| 2024-09-12 | 1.55294% | 1.4% | 2.78571% | 6.74286% |
| 2024-09-13 | 1.69412% | 1.5% | 2.94286% | 7.27143% |
| 2024-09-14 | 1.83529% | 1.6% | 3.1% | 7.8% |
| 2024-09-15 | 1.97647% | 1.67143% | 3.38571% | 8.38571% |
| 2024-09-16 | 2.11765% | 1.74286% | 3.67143% | 8.97143% |
| 2024-09-17 | 2.25882% | 1.81429% | 3.95714% | 9.55714% |
| 2024-09-18 | 2.4% | 1.88571% | 4.24286% | 10.1429% |
| 2024-09-19 | 2.54118% | 1.95714% | 4.52857% | 10.7286% |
| 2024-09-20 | 2.68235% | 2.02857% | 4.81429% | 11.3143% |
| 2024-09-21 | 2.82353% | 2.1% | 5.1% | 11.9% |
| 2024-09-22 | 2.96471% | 2.21429% | 5.37143% | 12.3% |
| 2024-09-23 | 3.10588% | 2.32857% | 5.64286% | 12.7% |
| 2024-09-24 | 3.24706% | 2.44286% | 5.91429% | 13.1% |
| 2024-09-25 | 3.38824% | 2.55714% | 6.18571% | 13.5% |
| 2024-09-26 | 3.52941% | 2.67143% | 6.45714% | 13.9% |
| 2024-09-27 | 3.67059% | 2.78571% | 6.72857% | 14.3% |
| 2024-09-28 | 3.81176% | 2.9% | 7% | 14.7% |
| 2024-09-29 | 3.95294% | 3.11429% | 7.22857% | 15.2714% |
| 2024-09-30 | 4.09412% | 3.32857% | 7.45714% | 15.8429% |
| 2024-10-01 | 4.23529% | 3.54286% | 7.68571% | 16.4143% |
| 2024-10-02 | 4.37647% | 3.75714% | 7.91429% | 16.9857% |
| 2024-10-03 | 4.51765% | 3.97143% | 8.14286% | 17.5571% |
| 2024-10-04 | 4.65882% | 4.18571% | 8.37143% | 18.1286% |
| 2024-10-05 | 4.8% | 4.4% | 8.6% | 18.7% |
| 2024-10-06 | 4.85714% | 4.5% | 8.84286% | 19.2571% |
| 2024-10-07 | 4.91429% | 4.6% | 9.08571% | 19.8143% |
| 2024-10-08 | 4.97143% | 4.7% | 9.32857% | 20.3714% |
| 2024-10-09 | 5.02857% | 4.8% | 9.57143% | 20.9286% |
| 2024-10-10 | 5.08571% | 4.9% | 9.81429% | 21.4857% |
| 2024-10-11 | 5.14286% | 5% | 10.0571% | 22.0429% |
| 2024-10-12 | 5.2% | 5.1% | 10.3% | 22.6% |
| 2024-10-13 | 5.57143% | 5.27143% | 10.5% | 23.0857% |
| 2024-10-14 | 5.94286% | 5.44286% | 10.7% | 23.5714% |
| 2024-10-15 | 6.31429% | 5.61429% | 10.9% | 24.0571% |
| 2024-10-16 | 6.68571% | 5.78571% | 11.1% | 24.5429% |
| 2024-10-17 | 7.05714% | 5.95714% | 11.3% | 25.0286% |
| 2024-10-18 | 7.42857% | 6.12857% | 11.5% | 25.5143% |
| 2024-10-19 | 7.8% | 6.3% | 11.7% | 26% |
| 2024-10-20 | 7.82857% | 6.4% | 12.0571% | 26.2143% |
| 2024-10-21 | 7.85714% | 6.5% | 12.4143% | 26.4286% |
| 2024-10-22 | 7.88571% | 6.6% | 12.7714% | 26.6429% |
| 2024-10-23 | 7.91429% | 6.7% | 13.1286% | 26.8571% |
| 2024-10-24 | 7.94286% | 6.8% | 13.4857% | 27.0714% |
| 2024-10-25 | 7.97143% | 6.9% | 13.8429% | 27.2857% |
| 2024-10-26 | 8% | 7% | 14.2% | 27.5% |
| 2024-10-27 | 8.21429% | 7.27143% | 14.5% | 28.2429% |
| 2024-10-28 | 8.42857% | 7.54286% | 14.8% | 28.9857% |
| 2024-10-29 | 8.64286% | 7.81429% | 15.1% | 29.7286% |
| 2024-10-30 | 8.85714% | 8.08571% | 15.4% | 30.4714% |
| 2024-10-31 | 9.07143% | 8.35714% | 15.7% | 31.2143% |
| 2024-11-01 | 9.28571% | 8.62857% | 16% | 31.9571% |
| 2024-11-02 | 9.5% | 8.9% | 16.3% | 32.7% |
| 2024-11-03 | 9.62857% | 8.94286% | 16.4571% | 33.0286% |
| 2024-11-04 | 9.75714% | 8.98571% | 16.6143% | 33.3571% |
| 2024-11-05 | 9.88571% | 9.02857% | 16.7714% | 33.6857% |
| 2024-11-06 | 10.0143% | 9.07143% | 16.9286% | 34.0143% |
| 2024-11-07 | 10.1429% | 9.11429% | 17.0857% | 34.3429% |
| 2024-11-08 | 10.2714% | 9.15714% | 17.2429% | 34.6714% |
| 2024-11-09 | 10.4% | 9.2% | 17.4% | 35% |
| 2024-11-10 | 10.4% | 9.27143% | 17.5286% | 35.2% |
| 2024-11-11 | 10.4% | 9.34286% | 17.6571% | 35.4% |
| 2024-11-12 | 10.4% | 9.41429% | 17.7857% | 35.6% |
| 2024-11-13 | 10.4% | 9.48571% | 17.9143% | 35.8% |
| 2024-11-14 | 10.4% | 9.55714% | 18.0429% | 36% |
| 2024-11-15 | 10.4% | 9.62857% | 18.1714% | 36.2% |
| 2024-11-16 | 10.4% | 9.7% | 18.3% | 36.4% |
| 2024-11-17 | 10.4% | 9.82857% | 18.5143% | 36.6% |
| 2024-11-18 | 10.4% | 9.95714% | 18.7286% | 36.8% |
| 2024-11-19 | 10.4% | 10.0857% | 18.9429% | 37% |
| 2024-11-20 | 10.4% | 10.2143% | 19.1571% | 37.2% |
| 2024-11-21 | 10.4% | 10.3429% | 19.3714% | 37.4% |
| 2024-11-22 | 10.4% | 10.4714% | 19.5857% | 37.6% |
| 2024-11-23 | 10.4% | 10.6% | 19.8% | 37.8% |
| 2024-11-24 | 10.4571% | 10.6429% | 19.8714% | 37.8286% |
| 2024-11-25 | 10.5143% | 10.6857% | 19.9429% | 37.8571% |
| 2024-11-26 | 10.5714% | 10.7286% | 20.0143% | 37.8857% |
| 2024-11-27 | 10.6286% | 10.7714% | 20.0857% | 37.9143% |
| 2024-11-28 | 10.6857% | 10.8143% | 20.1571% | 37.9429% |
| 2024-11-29 | 10.7429% | 10.8571% | 20.2286% | 37.9714% |
| 2024-11-30 | 10.8% | 10.9% | 20.3% | 38% |
| 2024-12-01 | 11% | 11.0143% | 20.4286% | 38.3143% |
| 2024-12-02 | 11.2% | 11.1286% | 20.5571% | 38.6286% |
| 2024-12-03 | 11.4% | 11.2429% | 20.6857% | 38.9429% |
| 2024-12-04 | 11.6% | 11.3571% | 20.8143% | 39.2571% |
| 2024-12-05 | 11.8% | 11.4714% | 20.9429% | 39.5714% |
| 2024-12-06 | 12% | 11.5857% | 21.0714% | 39.8857% |
| 2024-12-07 | 12.2% | 11.7% | 21.2% | 40.2% |
| 2024-12-08 | 12.2% | 11.7143% | 21.3143% | 40.3286% |
| 2024-12-09 | 12.2% | 11.7286% | 21.4286% | 40.4571% |
| 2024-12-10 | 12.2% | 11.7429% | 21.5429% | 40.5857% |
| 2024-12-11 | 12.2% | 11.7571% | 21.6571% | 40.7143% |
| 2024-12-12 | 12.2% | 11.7714% | 21.7714% | 40.8429% |
| 2024-12-13 | 12.2% | 11.7857% | 21.8857% | 40.9714% |
| 2024-12-14 | 12.2% | 11.8% | 22% | 41.1% |
| 2024-12-15 | 12.2714% | 11.8429% | 22.0286% | 41.2% |
| 2024-12-16 | 12.3429% | 11.8857% | 22.0571% | 41.3% |
| 2024-12-17 | 12.4143% | 11.9286% | 22.0857% | 41.4% |
| 2024-12-18 | 12.4857% | 11.9714% | 22.1143% | 41.5% |
| 2024-12-19 | 12.5571% | 12.0143% | 22.1429% | 41.6% |
| 2024-12-20 | 12.6286% | 12.0571% | 22.1714% | 41.7% |
| 2024-12-21 | 12.7% | 12.1% | 22.2% | 41.8% |
| 2024-12-22 | 12.7429% | 12.1% | 22.2429% | 41.8429% |
| 2024-12-23 | 12.7857% | 12.1% | 22.2857% | 41.8857% |
| 2024-12-24 | 12.8286% | 12.1% | 22.3286% | 41.9286% |
| 2024-12-25 | 12.8714% | 12.1% | 22.3714% | 41.9714% |
| 2024-12-26 | 12.9143% | 12.1% | 22.4143% | 42.0143% |
| 2024-12-27 | 12.9571% | 12.1% | 22.4571% | 42.0571% |
| 2024-12-28 | 13% | 12.1% | 22.5% | 42.1% |
| 2024-12-29 | 13.1143% | 12.2286% | 22.5857% | 42.1286% |
| 2024-12-30 | 13.2286% | 12.3571% | 22.6714% | 42.1571% |
| 2024-12-31 | 13.3429% | 12.4857% | 22.7571% | 42.1857% |
| 2025-01-01 | 13.4571% | 12.6143% | 22.8429% | 42.2143% |
| 2025-01-02 | 13.5714% | 12.7429% | 22.9286% | 42.2429% |
| 2025-01-03 | 13.6857% | 12.8714% | 23.0143% | 42.2714% |
| 2025-01-04 | 13.8% | 13% | 23.1% | 42.3% |
| 2025-01-05 | 13.8143% | 13% | 23.1143% | 42.4286% |
| 2025-01-06 | 13.8286% | 13% | 23.1286% | 42.5571% |
| 2025-01-07 | 13.8429% | 13% | 23.1429% | 42.6857% |
| 2025-01-08 | 13.8571% | 13% | 23.1571% | 42.8143% |
| 2025-01-09 | 13.8714% | 13% | 23.1714% | 42.9429% |
| 2025-01-10 | 13.8857% | 13% | 23.1857% | 43.0714% |
| 2025-01-11 | 13.9% | 13% | 23.2% | 43.2% |
| 2025-01-12 | 13.9143% | 13.0286% | 23.2286% | 43.2% |
| 2025-01-13 | 13.9286% | 13.0571% | 23.2571% | 43.2% |
| 2025-01-14 | 13.9429% | 13.0857% | 23.2857% | 43.2% |
| 2025-01-15 | 13.9571% | 13.1143% | 23.3143% | 43.2% |
| 2025-01-16 | 13.9714% | 13.1429% | 23.3429% | 43.2% |
| 2025-01-17 | 13.9857% | 13.1714% | 23.3714% | 43.2% |
| 2025-01-18 | 14% | 13.2% | 23.4% | 43.2% |
| 2025-01-19 | 14.0286% | 13.2% | 23.4% | 43.2% |
| 2025-01-20 | 14.0571% | 13.2% | 23.4% | 43.2% |
| 2025-01-21 | 14.0857% | 13.2% | 23.4% | 43.2% |
| 2025-01-22 | 14.1143% | 13.2% | 23.4% | 43.2% |
| 2025-01-23 | 14.1429% | 13.2% | 23.4% | 43.2% |
| 2025-01-24 | 14.1714% | 13.2% | 23.4% | 43.2% |
| 2025-01-25 | 14.2% | 13.2% | 23.4% | 43.2% |
| 2025-01-26 | 14.2429% | 13.2714% | 23.5% | 43.3286% |
| 2025-01-27 | 14.2857% | 13.3429% | 23.6% | 43.4571% |
| 2025-01-28 | 14.3286% | 13.4143% | 23.7% | 43.5857% |
| 2025-01-29 | 14.3714% | 13.4857% | 23.8% | 43.7143% |
| 2025-01-30 | 14.4143% | 13.5571% | 23.9% | 43.8429% |
| 2025-01-31 | 14.4571% | 13.6286% | 24% | 43.9714% |
| 2025-02-01 | 14.5% | 13.7% | 24.1% | 44.1% |
| 2025-02-02 | 14.5% | 13.7571% | 24.1% | 44.1143% |
| 2025-02-03 | 14.5% | 13.8143% | 24.1% | 44.1286% |
| 2025-02-04 | 14.5% | 13.8714% | 24.1% | 44.1429% |
| 2025-02-05 | 14.5% | 13.9286% | 24.1% | 44.1571% |
| 2025-02-06 | 14.5% | 13.9857% | 24.1% | 44.1714% |
| 2025-02-07 | 14.5% | 14.0429% | 24.1% | 44.1857% |
| 2025-02-08 | 14.5% | 14.1% | 24.1% | 44.2% |
| 2025-02-09 | 14.5286% | 14.1% | 24.2% | 44.2286% |
| 2025-02-10 | 14.5571% | 14.1% | 24.3% | 44.2571% |
| 2025-02-11 | 14.5857% | 14.1% | 24.4% | 44.2857% |
| 2025-02-12 | 14.6143% | 14.1% | 24.5% | 44.3143% |
| 2025-02-13 | 14.6429% | 14.1% | 24.6% | 44.3429% |
| 2025-02-14 | 14.6714% | 14.1% | 24.7% | 44.3714% |
| 2025-02-15 | 14.7% | 14.1% | 24.8% | 44.4% |

Table 5. Vaccine coverage rates for deterministic sensitivity analyses^2,11,12^

|  | **Upper Bound** | | | | **Lower Bound** | | | |
| --- | --- | --- | --- | --- | --- | --- | --- | --- |
| **Age group** | **12-17 years** | **18-49 years** | **50-64 years** | **65-100 years** | **12-17 years** | **18-49 years** | **50-64 years** | **65-100 years** |
| Annual | 22.0% | 21.1% | 37.2% | 66.5% | 9.8% | 9.4% | 16.6% | 29.7% |
| Sept | 6.1% | 4.9% | 11.2% | 23.7% | 2.7% | 2.2% | 5.0% | 10.6% |
| Oct | 13.6% | 12.6% | 23.5% | 46.7% | 6.1% | 5.6% | 10.5% | 20.8% |
| Nov | 16.2% | 16.3% | 30.4% | 56.9% | 7.2% | 7.3% | 13.6% | 25.4% |
| Dec | 19.9% | 18.7% | 34.2% | 63.2% | 8.9% | 8.3% | 15.2% | 28.2% |
| Jan | 21.7% | 20.4% | 36.0% | 65.9% | 9.7% | 9.1% | 16.0% | 29.4% |
| Feb | 22.0% | 21.1% | 37.2% | 66.5% | 9.8% | 9.4% | 16.6% | 29.7% |
| March | 22.0% | 21.1% | 37.2% | 66.5% | 9.8% | 9.4% | 16.6% | 29.7% |
| April | 22.0% | 21.1% | 37.2% | 66.5% | 9.8% | 9.4% | 16.6% | 29.7% |
| May | 22.0% | 21.1% | 37.2% | 66.5% | 9.8% | 9.4% | 16.6% | 29.7% |
| June | 22.0% | 21.1% | 37.2% | 66.5% | 9.8% | 9.4% | 16.6% | 29.7% |
| July | 22.0% | 21.1% | 37.2% | 66.5% | 9.8% | 9.4% | 16.6% | 29.7% |
| August | 22.0% | 21.1% | 37.2% | 66.5% | 9.8% | 9.4% | 16.6% | 29.7% |

Table 6. Vaccine coverage rates for additional vaccine coverage rate scenarios

|  | **10% increase overall population (12 to 64 high-risk and 65 plus) (1-dose)** | | | | **10% increase 65 plus only (1-dose)** | | | |
| --- | --- | --- | --- | --- | --- | --- | --- | --- |
| **Age group** | **12-17 years** | **18-49 years** | **50-64 years** | **65-100 years** | **12-17 years** | **18-49 years** | **50-64 years** | **65-100 years** |
| Annual | 16.2% | 15.5% | 27.3% | 48.8% | 14.7% | 14.1% | 24.8% | 48.8% |
| Sept | 4.5% | 3.6% | 8.3% | 17.4% | 4.1% | 3.3% | 7.5% | 17.4% |
| Oct | 10.0% | 9.2% | 17.3% | 34.3% | 9.1% | 8.4% | 15.7% | 34.3% |
| Nov | 11.9% | 12.0% | 22.3% | 41.8% | 10.8% | 10.9% | 20.3% | 41.8% |
| Dec | 14.6% | 13.8% | 25.1% | 46.4% | 13.3% | 12.5% | 22.8% | 46.4% |
| Jan | 16.0% | 15.0% | 26.4% | 48.4% | 14.5% | 13.6% | 24.0% | 48.4% |
| Feb | 16.2% | 15.5% | 27.3% | 48.8% | 14.7% | 14.1% | 24.8% | 48.8% |
| March | 16.2% | 15.5% | 27.3% | 48.8% | 14.7% | 14.1% | 24.8% | 48.8% |
| April | 16.2% | 15.5% | 27.3% | 48.8% | 14.7% | 14.1% | 24.8% | 48.8% |
| May | 16.2% | 15.5% | 27.3% | 48.8% | 14.7% | 14.1% | 24.8% | 48.8% |
| June | 16.2% | 15.5% | 27.3% | 48.8% | 14.7% | 14.1% | 24.8% | 48.8% |
| July | 16.2% | 15.5% | 27.3% | 48.8% | 14.7% | 14.1% | 24.8% | 48.8% |
| August | 16.2% | 15.5% | 27.3% | 48.8% | 14.7% | 14.1% | 24.8% | 48.8% |

### 4 Probability of Hospitalization and Outpatient Care

#### 4.1 Derivation of Probabilities (General Population)

The model requires the probability of hospitalization in those who do not receive annual vaccination (i.e. independent of prior vaccination history) and have a symptomatic infection.

The only data that is available, comes from healthcare databases and reflects the probability of hospitalization given medically attended SARS-CoV-2 infections. It was therefore necessary to transform the data so that the denominator was all symptomatic infections and not just symptomatic, medically attended infections.

Two sources providing the number of hospitalizations among medically-attended SARS-CoV-2 infections were available. Both studies examined individuals who received the XBB.1.5 COVID-19 vaccine compared to those that did not. Data used to estimate the probability of hospitalization were based on those that did not receive an XBB.1.5 vaccine. Kopel et al., (2024)^13^ provided data on adults aged 18 years and over with COVID-19 as any diagnosis for hospitalization. The Optum’s de-identified Clinformatics® Data Mart Database^14^ also had data on any diagnosis for reason of hospitalization. A database analysis was conducted based on Optum’s de-identified Clinformatics® Data Mart Database July 2024 release (extract June 2024, with claims through May 2024). The analysis began with all patients in the Optum’s de-identified Clinformatics® Data Mart Database, and inclusion criteria were (1) at least one day of enrolment between September 12, 2023 to February 29, 2024, and (2) patients with a COVID diagnosis as the primary diagnosis between September 12, 2023 and February 29, 2024. Patients with a missing age on the index date, defined as the date of first COVID diagnosis (in primary position) were excluded. COVID-19 diagnosis was identified using ICD-10-DX codes U071 COVID-19 or J1282 Pneumonia due to COVID. To identify ICU stays, additionally the following revenue codes were applied: 0200, 0201, 0202, 0203, 0204, 0206, 0207, 0208, 0209, 0210, 0211, 0212, 0213, 0214, 0219 OR ICU_IND = 'Y'. To identify inpatient stays with invasive mechanical ventilation, HCPCS/CPT* codes E0472 and K0534 as well as ICD-10-PCS codes 5A1935Z, 5A1945Z, and 5A1955Z were applied.

Because Kopel et al.^13^ is also used for the mRNA-1273 vaccine effectiveness data, it was selected as the primary source of data for hospitalization rates, with the Optum’s de-identified Clinformatics® Data Mart Database analysis used to estimate those under the age of 18 years. A number of steps were taken in order to derive the input for static model.

**Step 1:** The number of COVID-19 related hospitalizations were divided by the number of medically-attended COVID-19 cases.

Data were available for the three older age groups for Kopel et al., (2024) (i.e. for ages 18 years and above), and for all age groups from the Optum’s de-identified Clinformatics® Data Mart Database. As Kopel et al. did not include patients under the age of 18 years, the proportion hospitalized in this age group was estimated by taking the ratio of those aged 5-17 years to those aged 18-49 years from the Optum’s de-identified Clinformatics® Data Mart Database. This ratio was applied to the proportion hospitalized in those 18-49 years from Kopel. Values from step 1 are provided in Table 6.

Table 6. Proportion hospitalized in those medically attended

| **Age group** | **Proportion hospitalized** |
| --- | --- |
| 12-17 years | 3.69% |
| 18-49 years | 2.99% |
| 50-64 years | 4.46% |
| 65+ years | 13.49% |

**Step 2:** Calculate the proportion of COVID-19 related hospitalizations in symptomatic COVID-19 cases.

In order to estimate the proportion of all infections that were hospitalized, it was next necessary to transform the proportions in Table 6 to be the proportion hospitalized amongst all symptomatic infections. Age-specific data were not available on the proportion of individuals with symptomatic COVID-19 that seek medical attention. To obtain age-specific proportions of individuals who seek medical attention for their COVID-19 symptomatic illness, the CDC reported data on the proportion of individuals, by age group, with symptomatic influenza, that sought medical attention for their illness^15^ were considered as a starting point. In order to fit a separate dynamic transmission model (DTM) to the US population, these initial values were tested and subsequently varied during the calibration procedure of the DTM to achieve a better calibration fit. Particularly, the probability of seeking care was decreased in younger age groups to achieve a better calibration model fit of the SEIR model. The final values used for the base case calibration of the DTM SEIR model are displayed in Table 7. The values calculated in Step 1 were multiplied by the proportion of symptomatic individuals seeking medical attention in each corresponding age group to obtain the probability of hospitalization per symptomatic case (Table 8).

Table 7. Proportion of those with symptomatic COVID-19 seeking medical attention

| **Age group** | **Starting Point (CDC Influenza Estimates)**^15^ | **Final Proportions for Base Case** |
| --- | --- | --- |
| 0-17 years | 52.3% | 5.0% |
| 18-49 years | 37.6% | 9.5% |
| 50-64 years | 44.1% | 25.3% |
| ≥65 years | 65.1% | 61.5% |

CDC: Centers for Disease Control and Prevention

Table 8. Hospitalization probabilities given symptomatic infection

| **Age Group** | **Probability** |
| --- | --- |
| 0-17 years | 0.18% |
| 18-49 years | 0.28% |
| 50-64 years | 1.13% |
| 65+ years | 8.29% |

The final probabilities of hospitalization given symptomatic infection and proportion who do not seek care are summarized in the Table 9. The proportion seeking outpatient care is calculated as the remainder of 1 minus the probability of hospitalization and the proportion not seeking care. For ages 12 to 64 years, these probabilities were modified to reflect the fact that those considered high-risk have a higher probability of receiving hospitalization and outpatient care compared to the general population. The methods to make these adjustments are described in the next two sections below.

Table 9. General probability of hospitalization and proportion seeking care.

| **Age Group (Years)** | **Calculated: Hospitalization Probability in Those with Symptomatic Infection** | **Proportion Not Seeking Care** | **Proportion Seeking Outpatient Care** |
| --- | --- | --- | --- |
| 12 to 17 | 0.18% | 94.99% | 4.83% |
| 18-49 | 0.28% | 90.53% | 9.19% |
| 50-64 | 1.13% | 74.68% | 24.19% |
| ≥65 | 8.29% | 38.50% | 53.20% |

#### 4.2 Derivation Of Risk Ratios for High-Risk People (Outpatient and Hospital Care)

At the ACIP April 15/16^th^ 2025 meeting, results of an analysis were presented which estimated the prevalence of US adults having at least one CDC defined medical condition putting them at high risk of severe COVID-19^16^ to be 74%^17^. Within the same meeting, data on increased risk for COVID-19 hospitalization of US adults with underlying medical conditions were presented^18^. The presented adjusted rate ratios (RR) for COVID-19–associated hospitalizations among community-dwelling adults ages 18 years and older stratified by underlying condition and age showed an increased risk of COVID-19 patients having any underlying CDC-defined condition for COVID-19 hospitalization compared to the general population. For some conditions and ages, the adjusted RR was almost 10.

To parameterize the static health economic model for high-risk patients, however, these adjusted rate ratios could not be applied as the underlying the health economic model considers a probability of COVID-19 hospitalization given symptomatic COVID-19 infection derived for the general population. Instead of using these presented rate ratios, we followed the approach of Joshi et al. 2025^19^ and applied risk ratios of increased risk of COVID-19 outpatient attendance and COVID-19 hospitalization for those having a medical condition to the underlying probabilities of COVID-19 outpatient attendance and COVID-19 hospitalization given symptomatic infection which were applied in our age-based COVID-19 vaccination model.

The risk ratios for the high-risk population having at least one CDC defined high-risk medical condition were derived as follows:

1. Derivation of hazard ratios (HR) of increased risk of COVID-19 outpatient attendance and hospitalizations derived from Moderna real-world observational electronic health records and medical claims database analyses^20^
2. Transformation of the HRs into risk ratios (RR) using an optimal minimax transformation of hazard ratio (HR) using the formula described by VanderWeele, 2020^21^

##### 4.2.1 Step 1: Derivation of Hazard Ratios

The hazard ratios for increased risk of COVID-19 related outpatient attendance and hospitalization of those having chronic conditions or high-risk condition for severe COVID-19 (i.e., chronic kidney disease, immunocompromising conditions, cardiovascular disease, chronic lung disease and diabetes) were derived in an analysis using a widely used primary care electronic medical record (EMR) platforms in the US (i.e., the Veradigm EMR dataset, which includes the Allscripts Tier 1, Allscripts Tier 2, and Practice Fusion EMR) integrated with pharmacy and medical claims data (i.e., the Komodo dataset) with an observation period of March 15^th^ to December 15^th^, 2020. N=15,127,054 individuals were included in the analysis. Of these, n=9,418,745 individuals did not have any high-risk conditions (as defined above), n=3,604,418 had exactly one of these conditions, and n=2,103,891 had two or more of these conditions.

The estimated, adjusted HRs for COVID-19 related outpatient attendance and hospitalizations of those having exactly one of these conditions versus having none of these conditions were 1.175 (95% CI: 1.672-1.734) and 1.703 (95% CI: 1.672-1.734), respectively.

Despite these estimates being derived in the early phase of the COVID-19 pandemic (wild type SARS-Cov-2 circulating), more recent Omicron related evidence (such as data presented at the ACIP April 2025 meeting^18^ and the Moderna Bench to Practice webpage^22,23^) suggests an increased risk of patients with underlying medical conditions for severe COVID-19 by means of increased hospitalization rate ratios of these patients when compared to the general population or patients without those underlying conditions. Accordingly, we consider these estimates applicable for the analysis of mRNA-1283 COVID-19 vaccination in these high-risk patients.

##### 4.2.2 Step 2: Transformation of the HRs into RRs

The resulting risk ratios were estimated to be 1.17 (95% CI: 1,15, 1.78) for COVID-19 related outpatient visits and 1.69 (95% CI: 1.66, 1.72) for COVID-19 related hospitalizations.

#### 4.3 Application of Relative Risks to Derive High-Risk Inputs

Based on the derived risk ratios for increased risk of COVID-19 outpatient attendance and hospitalization of patients having a medical condition versus persons not having a medical condition, age-specific hospitalization probabilities given symptomatic infections in not at-high risk (i.e., those not having a medical condition) and high-risk population (i.e., those having a medical condition) were estimated as follows:

We define:

- H - age-specific hospitalization probability
- P – proportion
- _total – average population
- _non - not at-high risk population
- _high - high risk population

(a) H_total = (P_high * H_high) + (P_non * H_non)

It is shown that H_high = RR * H_non, hence the equation (a) could be rewritten as:

(b) H_total = (P_high * H_non * RR) + (P_non * H_non)

(c) H_total = H_non * (P_high * RR + P_non)

(d) H_non = (H_total) / (P_high * RR + P_non)

(e) H_high = RR * H_non

### 5 Mortality and Readmission

Mortality is assumed to affect hospitalized patients only (patients receiving no formal care and outpatient care are not subject to risk of death from COVID-19 infection). The age-specific probabilities of in-hospital mortality were adjusted for the high-risk analysis using a similar approach to estimating the probability of hospitalization given symptomatic infection. Joshi et al. (2025)^19^ adapted a static Markov model for high-risk adults to estimate the clinical and economic impact of vaccination strategies; high-risk adults were defined as those having been previously diagnosed with an immunocompromising condition, chronic lung disease, chronic kidney disease, cardiovascular disease, and diabetes mellitus. Joshi et al. estimate that approximately 29.3 million US adults have diabetes, and the CDC^24^ estimates that the prevalence of diabetes ranges from 9.4% to 13.1%. Joshi et al. estimated in-hospital mortality for patients with cardiovascular disease, diabetes mellitus, and immunocompromising conditions by applying relative risks to the COVID-19 general population mortality risk, resulting in relative risks of 1.23 for diabetes, 1.62 for cardiovascular disease, and 1.74 for immunocompromising conditions. Based on Joshi et al.^19^, it was conservatively assumed that the relative risk for in-hospital mortality for high-risk patients relative to non-high-risk patients was 1.23^25^ for all locations of care (i.e., no ICU or ventilator, ICU only, or ICU with ventilator), based on patients with diabetes.

The hospital readmission rate following discharge for COVID-19 and post-discharge mortality rate were obtained from a meta-analysis by Ramzi et al.^26^ Given that most hospital readmissions and post-discharge mortality occurred within the first 30 days post-discharge, the 30-day readmission rate estimated at 8.97%% (95% CI 8.37%-11.24%%) and the 30-day post-discharge mortality rate of 7.87% (95% CI 2.78%-12,96%%) were used in model analyses.^26^ These estimates were lower but comparable to the 1 year readmission and post-discharge rates estimates specific to the USA (10.0% and 8.1%, respectively).^26^ Further, the majority of COVID-19 hospitalizations in the US during 2023-2024 had an underlying medical condition and/or involved patients ≥65 years of age.^27^ Patients with underlying medical conditions and/or older age are at higher risk for all-cause readmission following hospitalization due to respiratory infections such as seasonal influenza^28,29^ or COVID-19.^26,29^ Finally, these estimates align with US quality metrics and are comparable with Omicron-specific estimates.^29,30^

The meta-analysis did not differentiate by age or in-hospital location of care and therefore readmission rates and post-discharge mortality were assumed to be the same for all ages and for the general ward, ICU only, and ICU with mechanical ventilation.

In their cost-effectiveness analysis of oral nirmatrelvir/ritonavir for high-risk COVID-19 patients, Carlson et al., (2024)^31^ applied an increased risk of post-discharge mortality to those who required mechanical ventilation. This was based on a study^32^ which found a 1.33 increased hazard ratio for mortality over 5 years in ICU patients compared to general ward patients. Although the original study was not specific to COVID-19, the possibility of increased post-discharge mortality was explored in a sensitivity analysis by increasing the rate by 33% to 10.5%.

Estimates of in-hospital mortality for the general population, stratified by age and in-hospital location of care, were obtained from the University of Michigan COVID-19 Vaccination Modeling Team and are based on COVID-NET surveillance data (March 2022 – October 2023). ^33^ Age-specific probabilities of in-hospital mortality were adjusted for the high-risk analysis using a similar approach to estimating the probability of hospitalization given symptomatic infection. Based on Joshi et al. (2025) ^19^, it was conservatively assumed that the relative risk for in-hospital mortality for high-risk patients relative to non-high-risk patients was 1.23, based on patients with diabetes.

Patients who survive the initial hospitalization are subject to risk of hospital readmission, and all patients who survive, including those who were readmitted, are subject to risk of post-discharge mortality. Unlike influenza surveillance systems, where death certificate data with pneumonia or influenza, other respiratory and circulatory causes, or other non-respiratory, non-circulatory causes of death are used to estimate deaths that occur outside the hospital (i.e., following discharge and capturing deaths related to readmission), similar surveillance systems for COVID-19 have not yet been developed. Accordingly, separate estimates for hospital readmission and post-discharge mortality related to COVID-19 are used in model analyses. The hospital readmission rate following discharge for COVID-19 and post-discharge mortality rate for the general population were obtained from a meta-analysis by Ramzi et al.^26^ Given that most hospital readmissions and post-discharge mortality occurred within the first 30 days post-discharge, the 30-day readmission (8.97%) and post-discharge mortality rates (7.87%) were used for the general population. Hospital readmission rates for high-risk patients were estimated using a similar approach as for the percentage of patients requiring hospitalization and the in-hospital mortality estimates. The RR for readmission for high-risk patients (RR=1.27) was obtained from Joshi et al. (2025) ^19^ based on the underlying risk estimate of Verna et al. (2021) ^25^ for patients with diabetes. The readmission rate for high-risk patients was calculated to be 9.49%; a weighted average of the high-risk (9.49%) and general population (8.97%) estimates of 9.35% was used for all locations of care in model analyses. Post-discharge mortality was assumed to be the same for the high-risk population as the general population.

### 6 Adverse Events

It was assumed that patients receiving no vaccine would not experience adverse events. Grade 3 and 4 Local and Systemic adverse event (AE) rates for those ≥12 years for mRNA-1283 and mRNA-1273 were estimated from Moderna clinical trial data (NextCOVE)^34^. AE rates for BNT162b2 were assumed to be equivalent to mRNA-1273; however, no grade 4 AE for BNT162b2 was assumed. All vaccines are also associated with a risk of myocarditis/pericarditis^35^ and anaphylaxis. ^36^

Table 11. Vaccine-related adverse event rates

| **Probability** | **mRNA-1283** | **mRNA-1273** | **BNT162b2** | **Source** |
| --- | --- | --- | --- | --- |
| Grade 3 Local | 1.61% | 1.17% | 1.17% | NextCOVE trial data |
| Grade 4 Local | 0% | 0% | 0% |  |
| Grade 3 Systemic | 7.16% | 5.77% | 5.77% |  |
| Grade 4 Systemic | 0% | 0.02% | 0% |  |
| Anaphylaxis | 0.0005% | 0.0005% | 0.0005% | Klein et al. (2021) ^36^ |
| Myocarditis/Pericarditis^‡^ | 0.0018% | 0.0018% | 0.0018% | Shimabukuro et al (2022)^35^ |

‡Myocarditis/pericarditis applies to ages 18-49 only; AE rates for BNT162b2 assumed equal to mRNA-1273; no grade 4 systemic AE was assumed for BNT162b2

### 7 Infection-Related Myocarditis

All patients with COVID-19 infection are subject to risk of infection-induced myocarditis, which is applied as a toll. The baseline rate of myocarditis in patients without COVID-19 infection and the increased risk due to COVID-19 infection was obtained from the CDC. Boehmer et al. (2021)^5^ conducted a cohort study on patients with at least one hospital-based encounter (either outpatient or inpatient) during March 2020-January 2021. The risk of myocarditis in patients with COVID-19 were compared to patients without COVID-19. The authors calculated the adjusted myocarditis risk difference, by age, between patients with and without COVID-19. These values were used to estimate the excess risk of myocarditis due to COVID-19. Where age groups reported in the CDC report did not align with age groups used in the model, a weighted average of the risks from the CDC reported age groups was used.

Table 12. Risk of myocarditis following SARS-CoV-2 infection

| Probability | Base (Range) | Source |
| --- | --- | --- |
| 12-17 years | 0.1220% | Boehmer (2021) ^37^ |
| 18-49 years | 0.0793% |  |
| 50-64 years | 0.1370% |  |
| 65+ years | 0.1800% |  |

**Assumes 50% female for all ages

### 8 Details on Scenario Analyses

#### 8.1 Indirect Benefit

The model was used to calculate the difference in the number of infections prevented in the vaccinated scenario compared to the unvaccinated scenario.  This difference was multiplied by 0.45 to estimate the additional infections prevented due to the indirect effect and added to the total number of infections prevented.

#### 8.2 Interim 2024-2025 Incidence

The estimation of the 2024-25 interim incidence was conducted as described in Section 2.2. The hospitalization targets are shown in Table 13 while the assumed average VE is shown in Table 14. Coverage rates from 2024/25 described in section 3 were used. The estimated incidence amongst the unvaccinated is shown in Table 18.

Table 13. Target hospitalization rates per 100,000 for the calculation of the interim 2024-2025 incidence scenario^1^

| **Age Group (Years)** | **Sep-23** | **Oct-23** | **Nov-23** | **Dec-23** | **Jan-24** | **Feb-24** | **Mar-24** | **Apr-24** | **May-24** | **Jun-24*** | **Jul-24*** | **Aug-24*** |
| --- | --- | --- | --- | --- | --- | --- | --- | --- | --- | --- | --- | --- |
| 12-17 | 1.7 | 1.1 | 0.6 | 0.9 | 0.8 | 0.8 | 0.7 | 0.6 | 0.3 | 0.1 | 0.1 | 0.1 |
| 18-49 | 4.3 | 2.7 | 2.0 | 3.1 | 4.0 | 2.9 | 2.4 | 1.6 | 1.1 | 0.4 | 0.4 | 0.4 |
| 50-64 | 13.1 | 8.7 | 6.2 | 9.2 | 12.4 | 9.1 | 6.9 | 4.3 | 2.9 | 1.0 | 1.0 | 1.0 |
| ≥ 65 | 66.0 | 46.8 | 30.8 | 54.7 | 59.3 | 40.5 | 35.3 | 24.2 | 17.2 | 4.9 | 4.9 | 4.9 |

* The hospitalization rate for July and August was assumed to be the same aa June.

Table 14. Market mix VE used in the calculation of infection incidence in the interim 2024/2025 incidence scenario

| **Age Group** | **Infections** | | **Hospitalizations** | |
| --- | --- | --- | --- | --- |
|  | **Initial VE** | **Waning*** | **Initial VE** | **Waning*** |
| 12-17 years | 0.6242 | 0.0475 | 0.6242 | 0.0246 |
| 18-49 years | 0.4573 | 0.0475 | 0.5441 | 0.0246 |
| 50-64 years | 0.4573 | 0.0475 | 0.5441 | 0.0246 |
| 65-100 years | 0.4573 | 0.0475 | 0.5441 | 0.0246 |

Table 15. Base case incidence of symptomatic infection (no vaccination arm) for the static model (% infected)

| **Age group (years)** | **Sep-23** | **Oct-23** | **Nov-23** | **Dec-23** | **Jan-24** | **Feb-24** | **Mar-24** | **Apr-24** | **May-24** | **Jun-24** | **Jul-24** | **Aug-24** |
| --- | --- | --- | --- | --- | --- | --- | --- | --- | --- | --- | --- | --- |
| 12-17 | 0.94 | 0.63 | 0.35 | 0.53 | 0.47 | 0.47 | 0.41 | 0.35 | 0.18 | 0.06 | 0.06 | 0.06 |
| 18-49 | 1.54 | 1.00 | 0.75 | 1.16 | 1.51 | 1.09 | 0.90 | 0.60 | 0.41 | 0.15 | 0.15 | 0.15 |
| 50-64 | 1.21 | 0.84 | 0.61 | 0.91 | 1.23 | 0.90 | 0.68 | 0.42 | 0.28 | 0.10 | 0.10 | 0.10 |
| 65+ | 0.87 | 0.67 | 0.46 | 0.82 | 0.89 | 0.61 | 0.52 | 0.36 | 0.25 | 0.07 | 0.07 | 0.07 |

#### 8.3 Inputs for VE Scenario Analyses

Table 16. mRNA-1283 initial VE inputs used for scenario analyses

| **Scenario** | **12-17 years** | | **18-49 years** | | **50-64 years** | | **65+ years** | |
| --- | --- | --- | --- | --- | --- | --- | --- | --- |
|  | **Infection** | **Hospitalization** | **Infection** | **Hospitalization** | **Infection** | **Hospitalization** | **Infection** | **Hospitalization** |
| ICATT | 68.5% | 76.7% | 64.0% | 69.4% | 64.0% | 69.4% | 62.8% | 73.5% |
| rVE infection = rVE hospitalization | 68.5% | 68.5% | 56.6% | 58.6% | 56.6% | 58.6% | 60.7% | 63.0% |

### 9 Deterministic Sensitivity Analysis Results

Table 17. Deterministic Sensitivity Analysis Results (mRNA-1283 relative to no vaccination)

| **Model Parameter** | **Scenario** | **Symptomatic infections averted** | | **Hospitalizations averted** | | **Deaths averted** | |
| --- | --- | --- | --- | --- | --- | --- | --- |
|  |  | **Number** | **% Change from Base** | **Number** | **% Change from Base** | **Number** | **% Change from Base** |
| Percentage hospitalized | Lower bounds (-25% of base-case) | 2,855,920 | 0% | 128,428 | -24.99% | 16,768 | -24.99% |
| Percentage hospitalized | Upper bounds (+25% of base-case) | 2,855,208 | 0% | 213,974 | 24.98% | 27,936 | 24.98% |
| Percentage ICU & ICU with MV | Lower bounds (minimum based on COVID-NET data) | 2,855,670 | 0% | 171,217 | 0.01% | 20,678 | -7% |
|  | Upper bounds (maximum based on COVID-NET data) | 2,855,377 | 0% | 171,192 | -0.01% | 25,352 | 13% |
| In-hospital mortality | Lower bound (-10% of base-case) | 2,855,619 | 0% | 171,213 | 0.00% | 21,466 | -4% |
|  | Upper bounds (+10% of base-case) | 2,855,508 | 0% | 171,203 | 0.00% | 23,240 | 4% |
| COVID-19 Incidence | Lower bounds (-25% of base-case) | 2,141,940 | -25% | 128,428 | -25% | 16,768 | -25% |
|  | Upper bounds (+25% of base-case) | 3,569,010 | 25% | 213,974 | 25% | 27,936 | 25% |
| Initial VE (infection) | Lower bound based on 95% CI '12-17: 50.8% 18-49: 48.5% 50-64: 48.5% 65+: 50.8% | 2,190,892 | -23% | 171,208 | 0% | 22,353 | 0% |
| Initial VE (infection) | Upper bound based on 95% CI '12-17: 85.2% 18-49: 64.4% 50-64: 64.4% 65+: 70.0% | 3,495,359 | 22% | 171,229 | 0% | 22,355 | 0% |
| Initial VE (Hospitalization) | Lower bound based on 95% CI '12-17: 56.2% 18-49: 46.3% 50-64: 46.3% 65+: 53.3% | 2,855,734 | 0% | 136,033 | -21% | 17,764 | -21% |
| Initial VE (Hospitalization) | Upper bound based on 95% CI '12-17: 92.4% 18-49: 88.2% 50-64: 88.2% 65+: 89.9% | 2,855,341 | 0% | 217,081 | 27% | 28,324 | 27% |
| Waning (Infection) | Lower bound: 3.05% | 3,406,388 | 19% | 171,208 | 0% | 22,353 | 0% |
|  | Upper bound: 6.75% | 2,309,816 | -19% | 171,208 | 0% | 22,353 | 0% |
| Waning (hospitalization) | Lower bound: 1.37% | 2,855,526 | 0% | 184,328 | 8% | 24,065 | 8% |
|  | Upper bound: 3.87% | 2,855,612 | 0% | 154,346 | -10% | 20,152 | -10% |
| Vaccine coverage | Upper bound based on ratio of high-risk to all adults in 2023-2024 | 4,277,508 | 50% | 256,436 | 50% | 33,480 | 50% |
| Vaccine coverage | Lower bound based on UK data following narrowing of vaccine recommendations | 1,907,445 | -33% | 114,453 | -33% | 14,943 | -33% |

### 10 Scenario Analysis Results

Table 18. Scenario Analysis Results (mRNA-1283 relative to no vaccination)

| **Model Parameter** | **Range/Data Selections** | **Symptomatic Infections Averted** | | **Hospitalizations Averted** | | **Deaths Averted** | |
| --- | --- | --- | --- | --- | --- | --- | --- |
|  |  | **Number** | **% Change*** | **Number** | **% Change*** | **Number** | **% Change*** |
| Base-Case | | 2,855,564 | -- | 171,208 | -- | 22,353 | -- |
| Infection Incidence | Interim 2024-2025 Scenario | 1,332,060 | -53% | 76,889 | -55% | 10,052 | -55% |
| Vaccine Effectiveness | rVE against infection = rVE against hospitalization | 2,856,510 | 0% | 142,345 | -17% | 18,589 | -17% |
| Vaccine Effectiveness | VE Infection (ICATT Study) | 3,252,056 | 14% | 171,255 | 0.0% | 22,359 | 0.0% |
| Indirect Benefit | Indirect Benefit Included | 4,140,567 | 45% | 220,080 | 29% | 28,736 | 29% |
| Proportion High-Risk | Increased to 50% of 12-17 year-olds | 2,900,083 | 2% | 171,370 | 0.1% | 22,367 | 0.1% |
| Inpatient Mortality | Increased mortality (RR=1.62) for high-risk patients | 2,855,569 | 0% | 171,208 | 0% | 22,293 | -0.3% |
| Percentage hospitalized | Increased RR of hospitalization (RR=1.45) for high-risk | 2,855,568 | 0% | 170,778 | -0.3% | 22,306 | -0.2% |
| Percentage hospitalized | Decreased RR of hospitalization (RR=1.66) for high-risk | 2,855,564 | 0% | 171,204 | 0% | 22,353 | 0% |
| Percentage hospitalized | Increased RR of hospitalization (RR=1.72) for high-risk | 2,855,563 | 0% | 171,280 | 0% | 22,362 | 0% |

RR: relative risk; rVE: relative vaccine effectiveness

*From Base-Case Results

Table 19. Scenario Analysis Results; Increase in Vaccination Coverage Rates (mRNA-1283 relative to no vaccination)

| **Scenarios Vaccination Coverage Rate Increase** | **Symptomatic Infections Averted** | | **Hospitalizations Averted** | | **Deaths Averted** | |
| --- | --- | --- | --- | --- | --- | --- |
|  | **Number** | **% Change*** | **Number** | **% Change*** | **Number** | **% Change*** |
| Base-Case | 2,855,564 | -- | 171,208 | -- | 22,353 | -- |
| 10% increase overall population (12 to 64 high-risk and 65 plus) (1-dose) | 3,141,120 | +10.0% | 188,329 | +10.0% | 24,588 | +10.0% |
| 10% increase 65 plus only (1-dose) | 2,972,331 | +4.1% | 186,438 | +8.9% | 24,374 | +9.0% |

*From Base-Case Results

14.	Moderna data on file 2024. Optum’s de-identified Clinformatics® Data Mart Database. Analysis September 2023 to February 2024 COVID-19 medical attendances and hospitalizations.

20.	Moderna. Data on File. Disease burden in patients with Medical conditions. Moderna Bench to Practice. 2024. In.

1. The COVID-NET data was downloaded on March 23, 2025.
